# A non-randomized, open-label, dose-finding, first-in-human trial of combined cytotoxic and immune-stimulatory gene therapy for primary adult high-grade glioma: transgene expression persists up to 17 months post-vector injection

**DOI:** 10.1101/2022.11.04.22281950

**Authors:** Yoshie Umemura, Daniel Orringer, Larry Junck, Maria L. Varela, Molly E.J. West, Syed M. Faisal, Andrea Comba, Jason Heth, Oren Sagher, Denise Leung, Aaron Mammoser, Shawn Hervey-Jumper, Daniel Zamler, Viveka N. Yadav, Patrick Dunn, Wajd Al-Holou, Todd Hollon, Michelle M. Kim, Daniel R. Wahl, Sandra Camelo-Piragua, Andrew P. Lieberman, Sriram Venneti, Paul McKeever, Theodore Lawrence, Ryo Kurokawa, Karen Sagher, David Altshuler, Lili Zhao, Karin Muraszko, Maria G. Castro, Pedro R. Lowenstein

**Affiliations:** Department Of Neurology, The University of Michigan School of Medicine; Department Of Neurosurgery, The University of Michigan School of Medicine; Department Of Cell and Developmental Biology, The University of Michigan School of Medicine; Department of Biomedical Engineering, The University of Michigan School of Medicine; The Rogel Cancer Center, The University of Michigan School of Medicine; Department Of Radiation Oncology, The University of Michigan School of Medicine; Department Of Pathology, The University of Michigan School of Medicine; Department Of Radiology, The University of Michigan School of Medicine; Dept. of Biostatistics, The University of Michigan School of Public Health.

## Abstract

**Background:** High-grade gliomas are fatal with universally poor prognosis. Initiation of effective cancer immune responses requires functional immune cells, particularly afferent antigen-presenting cells, which are typically absent from the brain parenchyma. To overcome this limitation, two adenoviral vectors expressing HSV1-TK and Flt3L were combined to target human gliomas. This first-in-human trial assessed safety, cytotoxicity, and recruitment of immune cells to the brain, in support of a future phase 1b/2 clinical trial.

**Methods:** Treatment-naïve high-grade glioma adult patients received injections of adenoviral vectors expressing HSV1-TK and Flt3L into the tumor bed, following maximal safe resection, at six escalating doses ranging from a total of 1.1×10^10^ to a maximum of 2×10^11^ viral particles. This was followed by two 14-day courses of Valacyclovir and standard upfront chemoradiation. Key inclusion criteria were age between 18 to 75, KPS≥70, and treatment-naïve possible high-grade glioma amenable to gross total resection. Patients were consented pre-operatively, and definitive enrollment occurred intraoperatively upon pathology confirmation of malignant glioma.

**Findings:** The treatment was well-tolerated without dose-limiting toxicity in patients with high grade glioma (n=17) (including 3 of the Gliosarcoma variant), or Anaplastic Ependymoma (n=1). The maximal-tolerated dose was not reached. The median overall survival was 21.3 months (95%CI: 11.1, 26.1) compared to 14.6 months with standard-of-care, with seven patients surviving for >2 years, three patients surviving for >3 years, and one patient still alive 57 months after enrollment. Tissue from subsequent re-resections from eight subjects showed elevated markers for CD3^+^, CD8^+^ T cells, and plasmacytoid dendritic cells (pDCs), suggesting the potential stimulation of anti-glioma immunity. Additionally, we detected biological activity from both viral vectors: (i) an increase in serum levels of Flt3L two weeks after vector administration, and (ii) expression of HSV1-TK in neurons, astrocytes, and SOX2+ cells in brain tumor samples up to 17 months post-vector injection into the brain.

**Interpretation:** Use of two adenoviral vectors expressing HSV1-TK and Flt3L appears to be both safe and feasible. Promising evidence from multiplex immunocytochemical analyses shows the presence of the expected immune infiltration, i.e., pDCs, along with persistent vector expression lasting up to 17 months post-injection. Moreover, the two-year survival rate of 38.8% compared to 19.6% with standard-of-care is promising, suggesting that this approach warrants further investigation in a phase 1b/2 clinical trial.

**Funding:** Funded in part by Phase One Foundation, Los Angeles, CA, The Board of Governors at Cedars-Sinai Medical Center, and The Rogel Cancer Center at The University of Michigan; clinicaltrials.gov: NCT01811992.

## Introduction

High-grade gliomas (HGG), of astrocytic, oligodendroglial or ependymal variants, are the most common types of primary brain tumor in adults.^1^ HGG are associated with poor prognosis, in spite of standard therapy, leading to a median overall survival (OS) around 14-16 months despite aggressive treatment with surgical resection and chemoradiation.^2^ Gene therapy for malignant gliomas using viral vectors encoding a direct or conditional tumoricidal approach has been shown to be safe in humans, though it has not demonstrated clinically significant benefits to date in large scale clinical trials.^3, 4^ Important challenges of gene therapy remain the low efficacy of direct tumor cell killing *in situ* and limited tumor and brain distribution of the transgenic therapeutic products preventing elimination of infiltrating malignant cells located at various distances from the primary tumor site.^5–11^

Initiation of an effective immune response against cancer requires functional dendritic cells (DCs), which are absent from the non-inflamed central nervous system (CNS),^12^ possibly underlying the lack of anti HGG immune responses (Figure 1A). Treatment with FMS-like tyrosine kinase 3 ligand (Flt3L) injected into the tumor bed, a cytokine known to cause proliferation and differentiation of DCs, recruits DCs into the tumor microenvironment (TME).^13–18^ Herpes Simplex Virus type 1-thymidine kinase (HSV1-TK) is an enzyme that phosphorylates the prodrug Valacyclovir and converts it into a nucleotide analog which causes cell death of mitotically active cells such as HGG.^9, 11, 19–21^ By combining Flt3L and conditionally cytotoxic HSV1-TK, the infiltrating DCs recruited into the TME become exposed to endogenous glioma antigens (released by HSV1-TK dependent tumor cell cytotoxicity), and become activated by TLR2 agonists, such as High Mobility Group Box 1 (HMGB1); these DCs then become capable of inducing specific anti-glioma immune responses against residual malignant glioma cells that remain post-surgical excision (Figure 1B).^22–26^ In pre-clinical studies, this approach induced T cell cytotoxicity and immune memory, high therapeutic efficacy and eliminated a large percentage (i.e., 50-75%) of tumors in various mouse models of glioblastoma (GBM).^14, 27–31^ The safety of this dual vector treatment in humans has not been established previously, though its safety in rats, canines and marmosets has been tested earlier by us.^26, 32–34^ We hypothesize that the co-administration of Ad-hCMV-TK and Ad-hCMV-Flt3L viral vectors to the post-resection tumor bed followed by oral Valacyclovir can be safely combined with standard-of-care (SOC) chemoradiation in patients with newly diagnosed HGG, and will potentially stimulate an anti HGG immune response.

**Figure 1:**
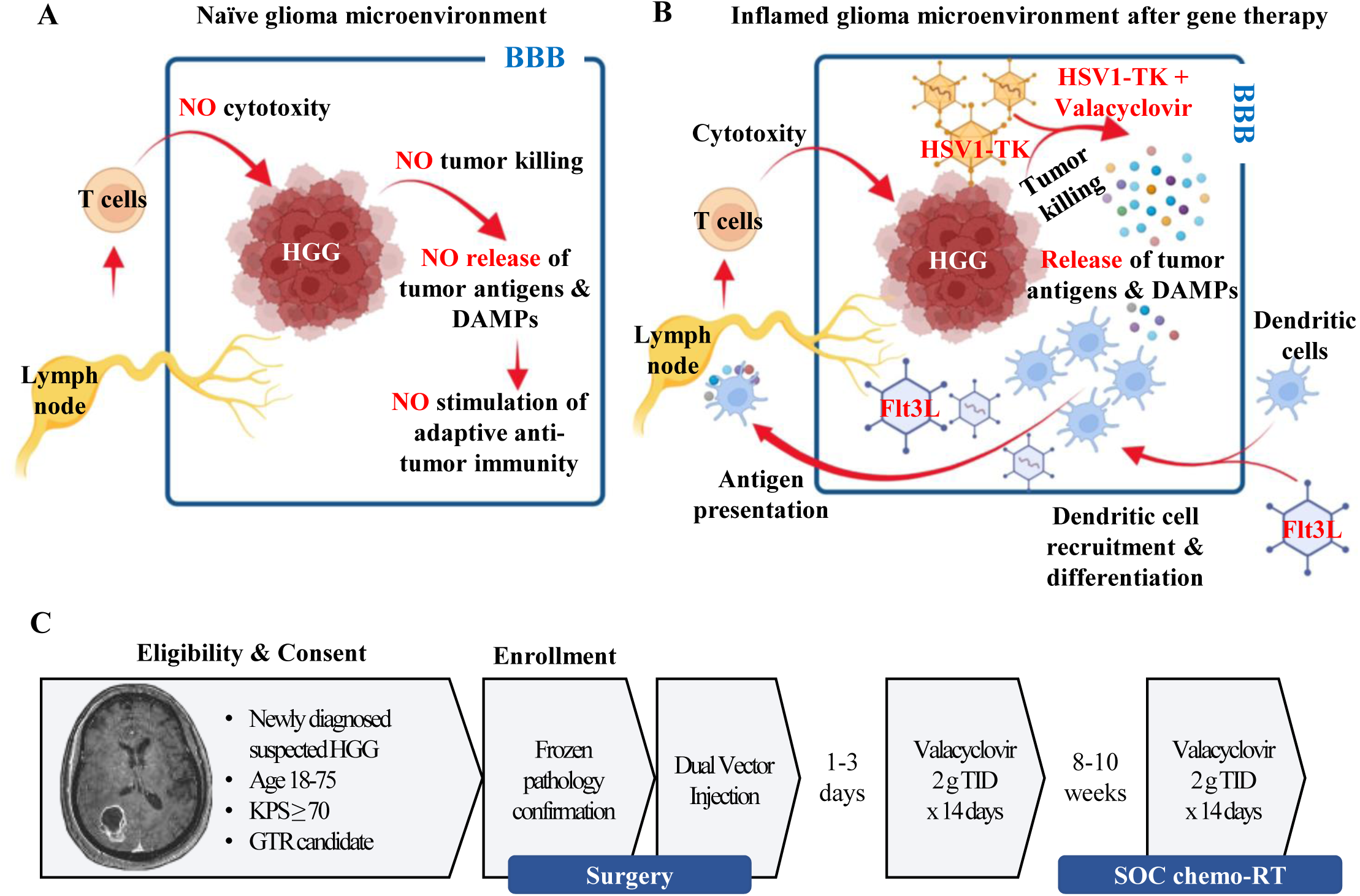
Overcoming lack of CNS dendritic cells by *in situ* immunotherapy using Flt3L and HSV1-TK. **A)** The central nervous system (CNS) lacks functional DCs, thus resulting in failure to initiate effective immune response against HGG. Blue border represents the blood brain barrier (BBB). **B)** Using *in situ* immunotherapy with Flt3L at the tumor bed, a cytokine known to cause proliferation of DCs, causes the migration of DCs into the TME. By combining Flt3L and HSV1-TK, a cytotoxic vector, DCs are recruited to choose tumor antigens within the glioma microenvironment and initiate systemic immunity, thus overcoming lack of CNS DCs. Blue border represents the blood brain barrier (BBB). **C)** In this study, adults with newly diagnosed HGG who underwent maximal safe resection received injection of HSV1-TK and Flt3L adenovectors to the tumor bed, followed by two 14-day cycles of Valacyclovir and SOC upfront chemoradiation.

## Panel: Research in Context

### Evidence before this study

We conducted a thorough search on PubMed to identify both preclinical and clinical studies related to the use of “TK AND Flt3L AND Glioma” as the search term. The search encompassed articles published between January 1st, 1993, and May 23rd, 2023. The results returned a total of 28 articles. The data surveyed strongly suggest that this particular approach holds promise in eliciting targeted immune responses against glioma in rats, dogs, and non-human primates (e.g., marmosets). These 13 articles were exclusively authored by our research group and primarily focused on pre-clinical models involving rodents, dogs, and primates. (Note: there is only one other team using Flt3L to treat pancreatic cancer –Hegde et al., Cancer Cell, 2020-, and one other group utilized a virus to express Flt3L in glioma in 2012 –Barnard et al., Neurosurgery, 2012-, yet never expanded that work further. We described the combined gene therapy much earlier in 2005 –Ali et al., Cancer Research, 2005). Work from supporting 28 papers studying the effects of TK and Flt3L in various models of gliomas served as the background information that was submitted to the FDA as the basis for the clinical evaluation of our approach in a phase 1 clinical trial (IND-14574; ClinicalTrials.gov: NCT01811992). The progress made in glioma treatment in recent years has been limited. Almost all clinical trials conducted thus far have failed to demonstrate clinical benefits for this devastating disease. Ad-TK has demonstrated the ability to induce infiltration of diverse immune cells, including macrophages and T cells, both in rodent models and clinical settings. However, the immune response against glioma by Ad-TK/GCV therapy often falls short without additional immunostimulatory strategies. By incorporating the co-expression of Flt3L, Ad-TK/GCV treatment stimulates a very potent antitumor immune response in experimental settings. This co-expression of Flt3L has been proven to attract dendritic cells (DCs) to the tumor microenvironment (TME), resulting in enhanced antigen presentation and subsequent infiltration and activation of T cells in glioma animal models. The safety and efficacy of the combined treatment has not been reported in human patients.

### Added value of this study

Brain tumors are poor stimulators of anti-tumor immune responses. We hypothesize that a lack of DCs within the brain explains the absence of immune responses against brain tumors. Therefore, stimulating dendritic cell infiltration will cause a robust anti-tumor immune response. To the best of our knowledge, this study represents the first-in-human trial designed to test dual-vector (Ad-hCMV-TK+Valacyclovir and Ad-hCMV-Flt3L) administration within the resection cavity walls following surgical resection. Our approach involves the administration of Ad-hCMV-TK, which expresses HSV1-TK, an enzyme that phosphorylates the prodrug Valacyclovir, leading to its cytotoxicity towards dividing tumor cells. We also administered Ad-hCMV-Flt3L, which expresses the cytokine Flt3L to recruit DCs to the brain and activate them. Upon killing of dividing tumor cells, the release of damage associated molecular patterns, such as HMGB1, triggers the activation of DCs through TLR2. Additionally, DCs take up tumor antigens. Consequently, activated DCs carrying tumor antigens migrate to lymph nodes to initiate a T-cell mediated anti-glioma immune response. Immunophenotyping of tumor biopsies obtained before dual vector administration revealed scarce infiltration of cytotoxic CD8+ cells, with myeloid cells and regulatory T cells (Tregs) being the most abundant immune populations. These findings are consistent with glioma induced immunosuppression. Paired biopsies obtained at first surgery and second surgery, i.e., following recurrence (after SOC treatment and administration of the combined gene therapy) were obtained from eight patients (Patient IDs #1, 2, 3, 5, 7, 8, 14, and 21). In these patients, we demonstrated evidence of immune infiltration by T cells and DCs using multiplex immunocytochemistry. Furthermore, our data provide evidence of the effective transduction and persistence of viral vector transgene expression, as immunoreactivity for HSV1-TK (expressed by Ad-hCMV-TK) was detected for up to 17 months post vector injection. We detected HSV1-TK immunoreactivity in 5/8 patients (Patient IDs #2, 7, 8, 14, and 21). Notably, a 38.8% two-year survival rate was observed (compared to the current records showing a 18% two-year survival rate), with one patient still alive nearly five years post-treatment. In addition, patients whose tumors express a mutated form of isocitrate dehydrogenase 1 (IDH1) live longer; none of the tumors in the patients in our trial expressed this mutated protein. These results support the further consideration of this approach in larger clinical trials.

### Implications of all the available evidence

This dual-vector gene therapy approach has demonstrated safety and feasibility in treating challenging high-grade gliomas (HGGs), which have historically shown resistance to conventional therapies. This strategy holds the potential of eliciting targeted anti-glioma immune responses against residual malignant glioma cells that persist after surgical resection. The long-term persistence of transgene expression (HSV1-TK) for up to 17 months post viral-vector administration suggests that continuous administration of Valacyclovir could augment the cytotoxic effect of Ad-hCMV-TK and lead to more favorable patient outcomes. Importantly, this dual-vector approach can be safely combined with SOC chemoradiation in patients with newly diagnosed HGGs. Our findings have the potential to improve overall survival and warrant further investigation in larger phase 2 clinical trials.

## Methods

### Objectives

Our primary objective was to identify the maximum tolerated dose (MTD) of Ad-hCMV-Flt3L and Ad-hCMV-TK, administered into the tumor bed after maximal safe resection of malignant gliomas. The secondary objective was to assess the potential benefit of Ad-hCMV-TK and Ad-hCMV-Flt3L dual-vector treatment of primary malignant glioma in OS at 12- and 24-months from intra-operative diagnosis. Exploratory objectives included the study of histological indices of inflammatory responses in tumor tissue from subsequent resections, when available, as well as measures of potential surrogate markers of intracranial immune responses. Viral vectors were produced at the Center for Cell and Gene Therapy, Baylor College of Medicine, Houston, TX. Examples of the last yearly quality control including the bioactivity of each vector is provided in the Supplemental Appendix [Figures S1 (Ad-hCMV-TK) and S2 (Ad-hCMV-Flt3L)].

### Patient Population

Adult patients with presumed newly diagnosed HGG who underwent maximal safe resection at the University of Michigan were potentially eligible. Key inclusion criteria were ages 18-75, KPS ≥70, and suspected newly diagnosed HGG amenable to gross total resection. Out of 21 subjects consented, two failed to meet eligibility criteria. In Cohort B one subject became unevaluable due to early withdrawal, not due to study-related toxicity, and was replaced to keep the total number of patients in Cohort B = 3. Eighteen subjects in all were evaluable and received study treatment. The eligibility section of the Supplemental Appendix contains detailed inclusion and exclusion criteria, and the CONSORT flow diagram is provided in Figure S3 of the Supplemental Appendix.

Eligible subjects who provided informed consent underwent maximal safe resection as indicated in the study schema (Figure 1C). When diagnosing a frozen brain tumor sample, the neuropathology group at The University of Michigan uses the term ‘malignant glioma’ and ‘high-grade glioma’ interchangeably. Importantly, this category includes GBM (WHO grade IV) and Anaplastic tumors of the astrocytic, oligodendroglial and ependymal lineage (WHO grade III tumors). As the trial enrolls patients upon intraoperative frozen diagnosis of ‘malignant glioma’, patients with Gliosarcoma and Anaplastic Ependymoma were included. Once the intra-operative pathology consultation confirmed HGG, subjects were enrolled in a phase 1, open label, dose escalation safety study of Ad-hCMV-TK and Ad-hCMV-Flt3L, assigned to six escalating doses in a 3+3 design (Supplemental Appendix Table S1, and Figure S3). The starting doses were Ad-hCMV-TK 1×10^10^ vp and Ad-hCMV-Flt3L 1×10^9^ vp (Cohort A), and the doses were escalated respectively to 1×10^11^ vp and 1×10^9^ vp (Cohort B), 1×10^10^ vp and 1×10^10^ vp (Cohort C), 1×10^11^ vp and 1×10^10^ vp (Cohort D), 1×10^10^ vp and 1×10^11^ vp (Cohort E), and finally, 1×10^11^ vp and 1×10^11^ vp (Cohort F). The starting dose of Ad-hCMV-TK was higher than Ad-hCMV-Flt3L because Ad-hCMV-TK has been utilized in various previous trials up to doses of 2×10^10, 12^ while ours was the first use of Ad-hCMV-Flt3L in humans.

The dual vectors were administered in a total volume of 2 ml to the tumor bed, divided into 0.1 ml injections at 20 sites. This was followed by two 14-day cycles of oral Valacyclovir 2g three times a day (TID) and SOC, upfront radiation and concurrent temozolomide per treating physician(s). Valacyclovir courses started at 1-3 days and 10-12 weeks after vector administration. This approach was based on our animal data showing viral transgene expression up to 6 and 12 months post-delivery of adenovirus into the brain.^35–37^ Dose modification for Valacyclovir was permitted if deemed necessary by the investigator and approved by the medical monitor, as outlined in the supplementary methods section of the Supplemental Appendix. Please refer to Table S2 for detailed information on the dose modification guidelines, as well as the renal adverse event management algorithms for Valacyclovir outlined in Table S3.

Depending on clinical status, Dexamethasone was administered prior to surgery and immediately following surgery at doses individualized to each patient’s clinical conditions. Doses typically start at 4-6 mg post-operatively every six hours. At various times after surgery the dose of Dexamethasone is tapered off depending on the individual patient’s condition with the aim of either maintaining lower doses of the steroid regimen, or attempting complete discontinuation.

This clinical trial was reviewed and approved by the Institutional Review Board-MED (University of Michigan School of Medicine) and registered at ClinicalTrials.gov (NCT01811992). An IND was obtained from the FDA prior to the study initiation (BB-IND 14574).

### Toxicity Evaluation

Toxicity was graded according to the Common Terminology Criteria for Adverse Events (CTCAE) version 4.0. Any one of the following occurring within 21 days of dual vector administration was considered a dose-limiting toxicity (DLT): 1) Grade 4 toxicity for constitutional symptoms with the exception of fever over 40°C for over 24 hours, 2) Grade 3 or higher neurologic toxicity relative to the changes from the pre-treatment neurological status and attributable to the study therapy regimen, 3) Grade 3 or higher non-hematologic toxicity attributable to the study therapy regimen, and 4) Grade 2 or higher autoimmune events.^9, 38, 39^ Constitutional symptoms considered as DLTs were not defined in the study protocol and are, unfortunately, not a term utilized in the Common Terminology Criteria for Adverse Events (CTCAE) in any specific section. According to the National Institutes of Health (NIH), however, constitutional symptoms refer to symptoms or manifestations indicating a systemic or general effect of a disease, potentially affecting the overall well-being or status of an individual (source: https://www.ncbi.nlm.nih.gov/medgen/3593). Further, in our protocol, it was not specified which adverse events from the CTCAE list would be considered constitutional symptoms. The determination of whether an adverse event qualified as a constitutional symptom was made by the treating clinical investigator based on the common definition of constitutional symptoms (such as the one provided by the NIH). Based on the protocol provided in the manuscript, hematologic toxicity was not considered a DLT, and hematological toxicity was not observed. Additional details in study design such as surgical, medical, and radiation protocols can be found in the Supplementary Methods section in the Supplemental Appendix.

### Immunohistochemistry on paraffin embedded brain biopsies

Resected tumor tissue from the primary resections, as well as available tissue from the second resections, were analyzed by immunohistochemistry (IHC), and by multiplex immunofluorescence.^22, 40–42^ Brain sections were subjected to endogenous peroxidase quenching carried out using a 0.3% H2O2 incubation for 5 minutes at room temperature, followed by heat-induced antigen retrieval, performed using 10 mM citric acid, 0.05% Tween 20, pH 6.0. Tissue permeabilization and blocking were completed using PBS 0.2% Tween-20 with 5% normal goat serum for one hour at room temperature. Tissue was then incubated with primary antibodies at 4°C overnight with concentrations as indicated: CD45 (Cell signaling #13917, 1:1000), CD3e (Cell signaling #85061, 1:400), CD11c (Cell signaling #45581, 1:1000), CD68 (Abcam ab955, 1:200), CD19 (Cell signaling #90176, 1:400), CD8 (Leica NCLL-CD8-4B11, 1:100), CD4 (Cell signaling # 48274, 1:500), HSV1-TK (In house, 1:1000), GFAP (Millipore #AB5541, 1:500), and SOX2 (ThermoFisher #MA5-31456, 1:1000). Sections for IHC-DAB detection were then incubated with biotinylated secondary antibodies at a 1:1000 dilution in PBS with 0.2% Tween-20 overnight at 4°C (Polyclonal Goat Anti-Rabbit Immunoglobulins/Biotinylated, Dako E0464, or Polyclonal Goat Anti Mouse Immunoglobulins/Biotinylated, Dako E0433). Avidin-Biotin-Complex (ABC) Binding reagent (Vectastain Elite ABC kit) and Betazoid DAB Chromogen detection kit (BioCare BDB2004) were used according to the manufacturer’s instructions. The IHC-stained slides were digitized at a single focal plane level at 40X magnification. Positive cells in the whole sections were quantified using QuPath v. 0.3.0. Sections for immunofluorescence detection were incubated with conjugated secondary antibodies at a dilution of 1:1000 Donkey anti-Rabbit IgG (H+L) highly cross-adsorbed secondary antibody, Alexa Fluor 555 (ThermoFisher #A-31572), Donkey anti-Chicken IgG (H+L) Alexa Fluor 647 (Jackson ImmunoResearch # 703-605-155), and Donkey anti-Mouse IgG (H+L) highly cross-adsorbed secondary antibody, Alexa Fluor 488 (ThermoFisher # A-21202). Details of the multiplex immunocytochemistry and statistical analysis are provided in the Supplementary Methods, in Correlative Studies within the Supplemental Appendix.

### HSV1-TK detection specificity

To determine the *in situ* specificity of HSV1-TK detection, we infected the human glioma cell line SJGBM with Ad-TK or Ad-βGal (MOI=100) and fixed the cells with PFA 48 hours after infection. We then incubated the antibody solution of HSV1-TK (1:1000) for 24 hours with fixed, permeabilized infected cells to obtain: (1) antibody solution pre-absorbed against cells infected with Ad-βGal, and (2) antibody solution pre-absorbed against cells infected with Ad-TK. We expected that antibody pre-absorbed with Ad-βGal would not recognize any elements from the viral infection, but would recognize the HSV1-TK antigen, while antibodies pre-absorbed against cells infected with Ad-TK would not recognize any HSV1-TK present in the human samples. We employed the pre-adsorbed antibody solutions for IHC using the DAB method, or immunofluorescence as described in the Immunohistochemistry on Paraffin-Embedded Brain Biopsies (IHC-DAB) section.

### Statistical Analysis

Demographic data was summarized in a tabular format. Kaplan-Meier method was used to estimate median, 12- and 24-months OS along with 95% confidence intervals. OS was defined from the time of study enrollment (the date of dual vector injection) to the time of death. Progression-free survival (PFS) was defined from the time of study enrollment to the time of first progression or death using iRANO criteria. RStudio Version 1.3.1073 was used for statistical analysis.

This trial is registered with ClinicalTrials.Gov, NCT01811992.

## Results

### Patients Characteristics

Eighteen patients with median age of 64 years (range 45-73) were assigned to six dose levels. As summarized in Table 1A, eight subjects were women (44%). The KPS at the time of enrollment was 90-100 in fifteen subjects (83%), 70-80 in three subjects (17%). In eleven participants (61%) radiographic gross total resection was achieved prior to the dual vector administration. All participants met the inclusion criteria of having the intraoperative pathology diagnosis of HGG. Final diagnosis was as follows: 17 tumors (95%) were diagnosed as GBM, WHO grade 4 (3 of them were of the Gliosarcoma subtype), and one was an Anaplastic Astrocytoma, WHO grade 3Glioblastoma (5%). All tumors were IDH wildtype, and 39% (n=7) had MGMT promoter methylation. Comprehensive information regarding the baseline demographic and clinical characteristics of each patient is provided in Table S4 of the Supplemental Appendix.

**TABLE 1A:** Patient Characteristics.

| Characteristic | Total N=18 |
| --- | --- |
| Median age (range) | 64 (45-73) |
| Women – no. (%) | 8 (44) |
| KPS – no. (%) |  |
| 90-100 | 15 (83) |
| 70-80 | 3 (17) |
| Radiographic GTR – no. (%) | 11 (61) |
| MGMT methylated – no. (%) | 7 (39) |
| **IDH mutation – no. (%)** | **0 (0)** |
| Final diagnosis – no. (%) |  |
| Glioblastoma | 14 (77.7) |
| Gliosarcoma | 3 (16.6) |
| Anaplastic Ependymoma | 1 (5.5) |

### Study Treatment Tolerability

Six subjects (33%) experienced adverse events that were at least possibly related to study treatment which ranged from CTCAE grades 1 to 3 (Table 1B) and consisted of one subject per following toxicities: dyspepsia (grade 1), elevated creatinine (grade 2), dizziness (grade 1), encephalopathy (grade 3), headache (grade 1), seizure (grade 2 & 3), confusion (grade 2), hallucinations (grade 1), acute kidney injury (grade 3), and maculo-papular rash (grade 1).

**TABLE 1B:** Adverse Events At Least Possibly Related to Study Treatment in Evaluable Patients.

| Event | Dual Vector Dose (N=3 per level) |  |  |  |  |  | Total |
| --- | --- | --- | --- | --- | --- | --- | --- |
|  | A | B | C | D | E | F | N=18 |
|  | *no.* |  |  |  |  |  | *no. (%)* |
| Gastrointestinal disorders |  |  |  |  |  |  |  |
| Dyspepsia, Grade 1 | - | - | - | 1 | - | - | 1 (6) |
| Investigations |  |  |  |  |  |  |  |
| Creatinine increased, Grade 2 | - | - | - | - | - | 1 | 1 (6) |
| Nervous system disorders |  |  |  |  |  |  |  |
| Dizziness, Grade 1 | - | - | - | - | - | 1 | 1 (6) |
| Encephalopathy, Grade 3 | - | - | - | - | - | 1* | 1 (6) |
| Headache, Grade 1 | - | - | - | 1 | - | - | 1 (6) |
| Seizure, Grade 2 | - | 1* | - | - | - | - | 1 (6) |
| Grade 3 | 1 | - | - | - | - | - | 1 (6) |
| Psychiatric disorders |  |  |  |  |  |  |  |
| Confusion, Grade 2 | - | - | - | - | - | 1 | 1 (6) |
| Hallucinations, Grade 1 | - | - | - | - | - | 1 | 1 (6) |
| Renal and urinary disorders |  |  |  |  |  |  |  |
| Acute kidney injury, Grade 3 | - | - | - | - | - | 1* | 1 (6) |
| Skin and subcutaneous disorders |  |  |  |  |  |  |  |
| Rash maculo-papular, Grade 1 | - | 1 | - | - | - | - | 1 (6) |
| Total no. patients with study related events | 1 | 2 | - | 2 | - | 1 | 6 (33) |

\*Serious adverse events

Ten subjects experienced serious adverse events (Table S5 of the Supplemental Appendix) which ranged from CTCAE grades 2 to 5. There was one grade 5 serious adverse event (SAE) observed in Patient ID #6. The SAE involved the development of pneumonia, which subsequently led to respiratory failure and, unfortunately, resulted in the patient’s death. It is important to note that this grade 5 toxicity was not considered to be related to the study treatment. Subject ID #15 experienced an intracranial hemorrhage within the tumor bed, which was surgically evacuated. Thus, it was given a grade 4 classification. Seven days following the initial surgery and dual vector treatment, the patient’s primary care physician initiated Enoxaparin treatment for prevention of pulmonary embolism/deep vein thrombosis. Six days after starting Enoxaparin, the patient presented with an intracranial hemorrhage and underwent surgical evacuation two days later. Subsequently, the patient was discharged in a stable condition three days after the evacuation. The occurrence of the intracranial hemorrhage was not considered to be related to the study treatment, but related to the anti-coagulant treatment.

One subject experienced three serious adverse events of grades 2-3, which were possibly related to study treatment. Serious adverse events possibly related to the study treatments were grade 2 seizure (n=1), grade 3 encephalopathy (n=1), and grade 3 acute kidney injury (n=1), all in the same patient, which were possibly related to Valacyclovir. No DLT of the dual viral vectors was observed, and the maximal tolerated dose of HSV-TK and Flt3L vectors was not reached. Other adverse events which were unrelated to study treatments are summarized in Table S6 of the Supplemental Appendix; (see also Supplementary Methods: Toxicities and Dose Delays).

### Upfront Treatment (see also Supplemental Appendix: Protocol Treatment Plan)

Study subjects received upfront radiation and concurrent temozolomide per SOC by treating oncologists. Detail of adjuvant treatment rendered was available for 17 subjects. Details of SOC chemoradiation for one subject who elected to receive care closer to home were unavailable. The one patient with Anaplastic Ependymoma received a total 59.4 Gy of radiotherapy without concurrent temozolomide followed by imaging surveillance without adjuvant chemotherapy.

Of 16 patients with GBM with known radiation details, all received concurrent temozolomide, 14 patients received the standard total 60 Gy, one patient received hypofractionated 40.5 Gy total, and one patient received 10 Gy before interruption due to wound issues and later completed 40.5 Gy between adjuvant temozolomide cycles. All 16 patients received SOC sequential adjuvant cyclic temozolomide (range 1-11 cycles total). One patient received bevacizumab shortly after completion of concurrent chemoradiation for disease burden from post-radiation treatment changes along with cyclic temozolomide. None of the subjects opted to use tumor treating fields as part of the upfront treatment.

### Progression-Free and Overall Survival

The median OS from the time of enrollment was 21.3 months (95%CI: 11.1, 26.1). The 12-month OS was 72% (95%CI: 45.6, 0.87) and the 24-month OS was 38.8% (95%CI: 17.5, 60.0). (Figure 2A). Notably, the three-year survival rate was 16.66% (3/18 patients), the four-year survival rate was 11.11% (2/18 patients), and one patient is still alive 57 months after enrollment (Patient ID #16). The median progression free survival was 9.9 months (95%CI: 7.3, 13.3). (See also Supplemental Appendix, page 10, Clinical and Imaging Evaluation)

**Figure 2:**
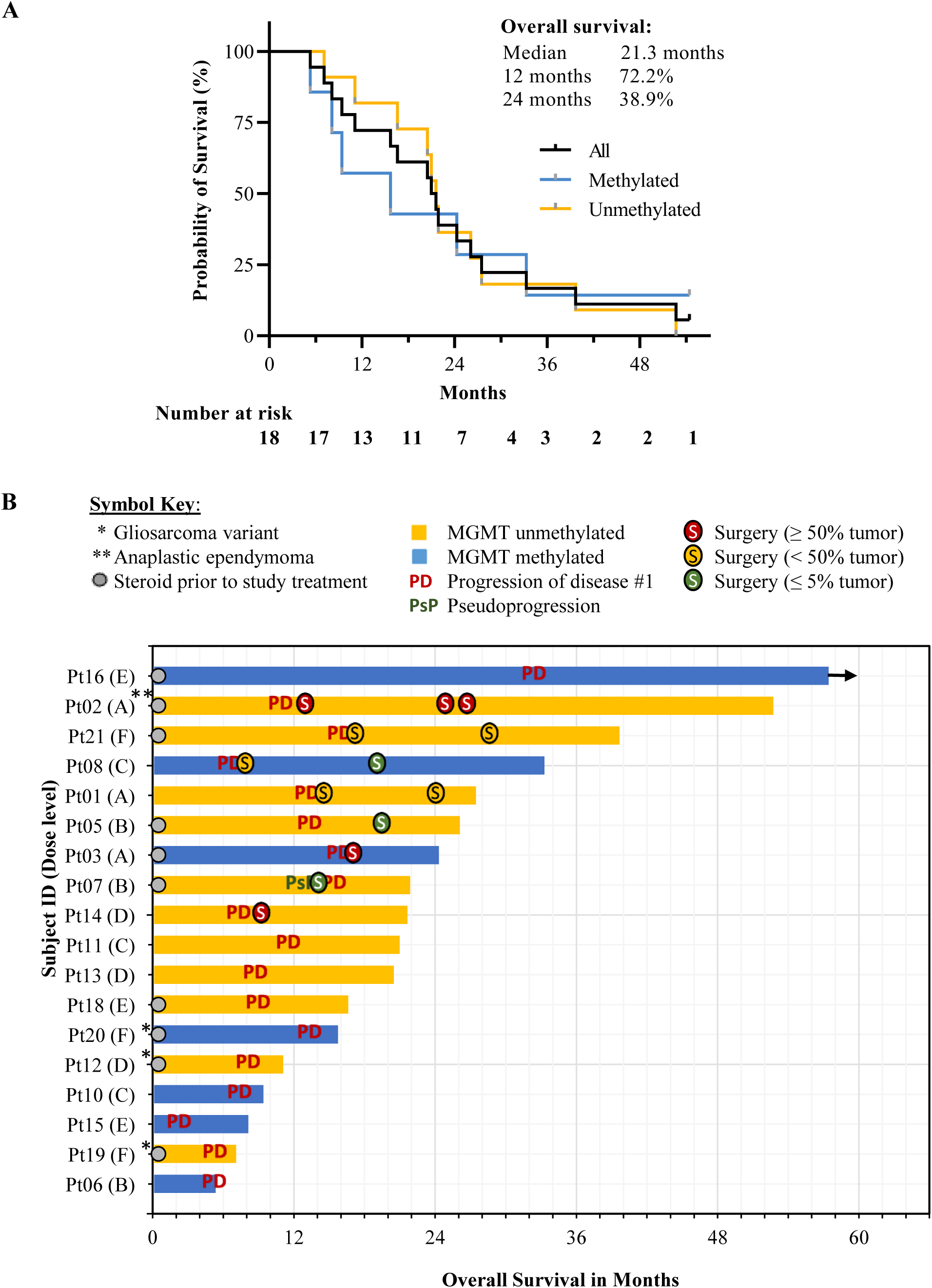
Dual viral vector approach shows encouraging survival outcomes. **A)** Using Kaplan-Meier method, median OS was 21.3 months. **B)** Swimmer plots of bars representing each study subject’s OS in months, noting those with steroid use at the time of enrollment. One patient with a rightward arrow is still alive and is currently followed for survival. Annotations correspond to respective timing of first progression and PsP per iRANO criteria (if known), and subsequent surgery with proportion of viable tumor noted in the sampled specimen. One patient had the diagnosis of Anaplastic Astrocytoma (**) and all other patients had the diagnosis of HGG, including three with the Gliosarcoma variant (*). All 18 subjects had the frozen pathology of malignant glioma prior to enrollment at the time of tumor resection.

Pre-operative diffusion-weighted imaging (DWI) (n=9) and dynamic susceptibility contrast-enhanced (DSC) (n=18) parameters were evaluated. There were weak positive correlations between baseline pre operative normalized relative cerebral blood volume (nrCBV) and OS, and between pre-operative normalized apparent diffusion coefficient (nADC) mean and OS (r = 0.53 and 0.49, respectively). No significant correlations were found between MRI parameters and survival (OS or PFS). MRI results are discussed in Supplementary Results within the Supplemental Appendix.

### Surgical Pathology at Suspected Recurrence

The timing of subsequent surgeries in relation to radiographic progression is illustrated in Figure 2B. Eight patients (44%) underwent at least one subsequent resection (Patient IDs #1, 2, 3, 5, 7, 8, 14, and 21). After the dual vector and Valacyclovir therapy and upfront SOC therapy, seven patients had re-operation at the time of radiographically suspected tumor progression; six subjects had surgically confirmed progression, though the resected tissues from three of these subjects had extensive treatment related inflammation and necrosis, and <50% was tumor; one subject (ID #7) had surgically confirmed pseudoprogression (PsP). PsP was defined as ≤ 5% proliferating tumor involvement of resected tissue, which was assessed using Ki67 staining. One other subject (ID #5) received second line therapy without surgical confirmation of first progression but underwent re-resection after the second line therapy, at which time PsP was noted.

When further progressions were suspected, four subjects (ID #1, 2, 8, 21) underwent additional resections; three had two re-resections (ID #1, 8, 21), and one subject had three re-resections (ID 2). Subject ID #1, 8, and 21 had surgical confirmation of progression after first-line therapy but only <50% tumor involvement of the resected tissue; subject ID #1 (Figure 3A) and 21 had similar findings at subsequent re-resection with majority of the resected tissue showing treatment related changes albeit confirmed progression (tumor >5%); subject ID #8 had surgically confirmed PsP after the second-line therapy of procarbazine and lomustine.

**Figure 3:**
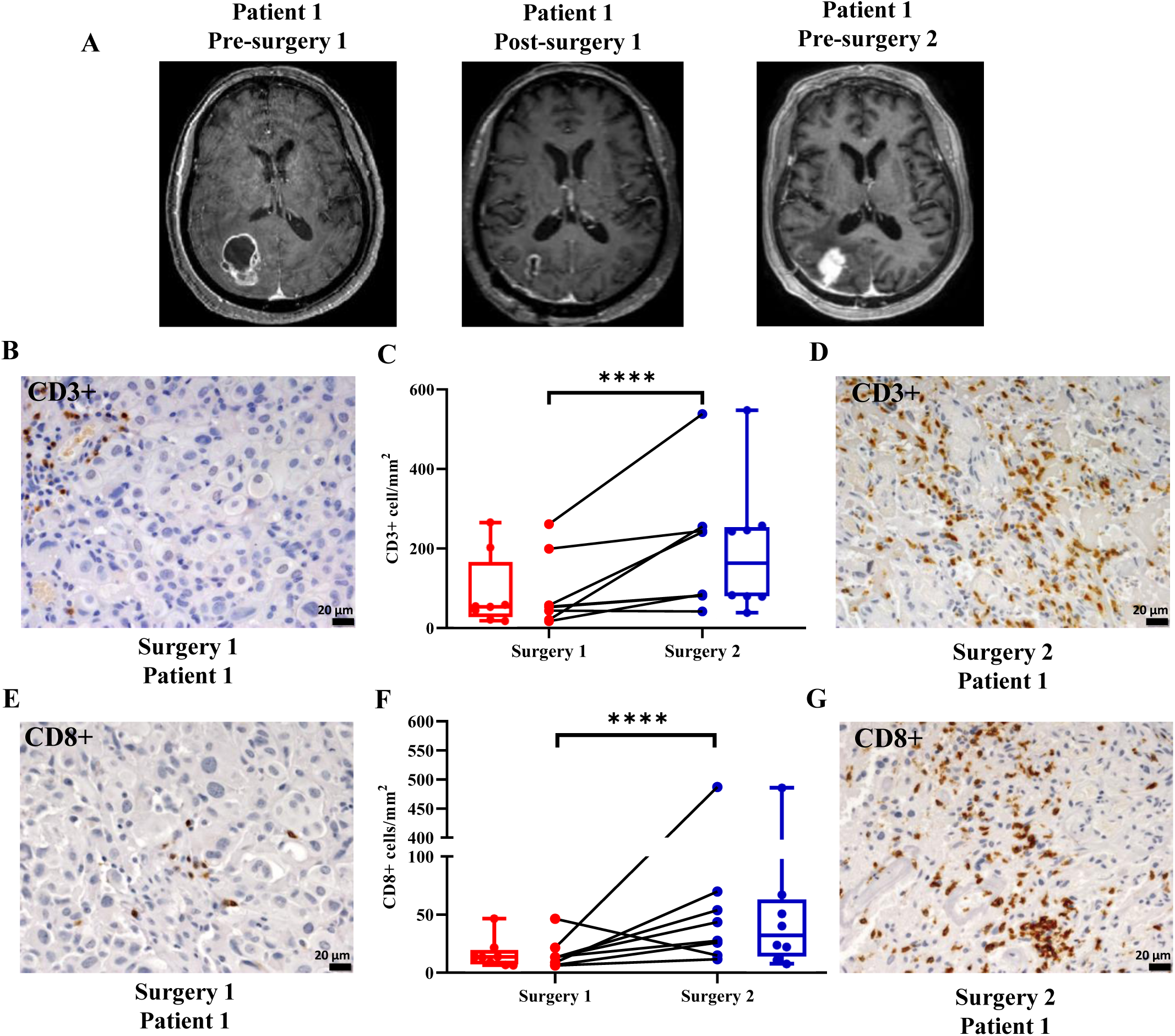
Immunohistochemical analysis of CD3+ and CD8+ cells in human HGG tissue at primary resection, pre-vector TK-Flt3L treatment, and at secondary surgery due to presumed tumor recurrence. **A)** Representative initial MRI scan of patient ID #1 before primary resection (pre-surgery 1), post-surgery 1 taken approximately 100 days after the initial MRI, and pre-surgery 2 at potential tumor recurrence, which occurred around 13 months and 5 days after the initial MRI. **B and D**) Representative images of CD3 expression were obtained; Scale bar: 20 μm (Patient ID #1). **C)** Box plots and lines graphs represent the quantification of CD3+ cells numbers (cells/mm^2^) using QuPath positive cell detection on HGGs samples obtained pre-TK-Flt3L treatment (Surgery 1) and post-surgery upon recurrence (Surgery 2). Error bars represent mean±SEM. The analysis included a total of 8 patients (Patient IDs #1, 2, 3, 5, 7, 8, 14, and 21), **** p<0.0001. **E and G**) Representative images of CD8 expression were obtained; Scale bar: 20 μm (Patient ID #1). **F)** Box plots and lines graphs represent the quantification of CD8+ cells numbers (cells/mm^2^) using QuPath positive cell detection on HGGs samples obtained pre-TK-Flt3L treatment (Surgery 1) and post-surgery upon recurrence (Surgery 2). Error bars represent mean±SEM. The analysis included a total of 8 patients (Patient IDs #1, 2, 3, 5, 7, 8, 14, and 21), and statistical analysis revealed a highly significant difference (**** p<0.0001) in both CD3+ and CD8+ cell numbers between Surgery 1 (pre-treatment) and Surgery 2 (post-treatment). For each marker, a linear mixed effects (LME) model was used to compare NumPos (log-transformed) between two surgery procedures. Random effect was included in the LME model to consider correlations between data measured on the same subject. Significance is determined if p<0.05. All analyses were conducted using SAS (version 9.4, SAS Institute, Cary, NC).

Subject ID #2 had the diagnosis of Anaplastic Ependymoma from the tissue obtained from the initial surgery prior to the dual vector injections, and the first surgically confirmed local recurrence showed recurrent/residual Anaplastic Ependymoma. Subsequently, this patient developed a contralateral distant lesion which was biopsied and showed HGG that was morphologically different from the original Anaplastic Ependymoma. This lesion was then resected two months later, which was notably more infiltrative than the original tumor at the initial diagnosis, with features of astrocytoma with areas of necrosis and microvascular proliferation consistent with GBM. It was unclear if this distant lesion was a new primary tumor or a dedifferentiation of the known Anaplastic Ependymoma.

### Immunocytochemistry Analyses

Immunocytochemical analysis of immune infiltrating cells was performed on tumor sections at primary resection, i.e., pre dual vector TK-Flt3L treatment, and compared with the same patient’s samples taken at secondary surgery due to potential tumor recurrence (n=8) (Patient IDs #1, 2, 3, 5, 7, 8, 14, and 21). There were no differences in: hematopoietic CD45 positive cells (Figure S4), or monocytic lineage cells (CD68 positive cells) (Figure S5).

Importantly, CD3e positive cells (T-lymphocytes) were significantly increased in the re-resection sample (surgery 2), in comparison with primary resection (surgery 1) (p<0.0001, Figure 3B-D). Deeper characterization of T-cells showed that there was no difference in CD4+ cells (Figure S6), while there was a statistically significant increment of CD8+ cells (p<0.0001, Figure 3E-G) in surgery 2 samples.

### Multiplex Immunocytochemistry Analysis

Multiplex immunocytochemical analysis of the TME was performed on tumor sections at primary resection, prior to administering adenoviral vectors expressing either HSV1-TK or Flt3L, and then compared with the same patient’s samples obtained during secondary surgery due to presumptive tumor recurrence (n=8).

### Neighborhood Analysis

After clustering neighborhoods (as defined in Supplementary Methods: Correlative Studies, within the Supplemental Appendix) with similar compositions into regions using CytoMAP, three distinct regions were identified in all tissue samples: the Immune Region, the Tumor Region, and the Other Region. The Immune Region, shown in gold in Figure 4A-C and in Supplemental Appendix (Figure S9 and Figure S10), was defined by a high density of CD45+ cells compared to the rest of the tissue. The Tumor Region, shown in teal in Figure 4A-C and Figure S9-S10, was defined by a high density of SOX2+ cells compared to the rest of the tissue. The Other Region, shown in purple in Figure 4A-B and Figure S9-S10, was characterized by a high density of cells that were both SOX2- and CD45-, making them impossible to phenotype using the scheme used. Across all patients analyzed (Patient IDs #1, 2, 3, 5, 7, 8, 14, and 21), the Tumor Region (enriched in SOX2+ cells) decreased in size by 36.5 ± 10.7% (range 3.6-47.4%; n=8). In contrast, the Immune Region (enriched in CD45+ cells) increased in six patients by 21.98 ± 5.6% (n=6), while it decreased in two patients by 18.1%, and 23.6%. On average, the Tumor Region accounted for a smaller proportion of the analyzed tissue, while the Immune Region expanded (Figure 4D). Analysis of region composition, or average number of cells per neighborhood within a region, showed that within the Tumor Region, the density of various immune cell types increased in samples obtained from the second surgical resection. These immune cell types included CD8+ T cells, double positive T cells, conventional DCs (cDCs)/monocytes, and pDCs (Figure 4F-G).

**Figure 4:**
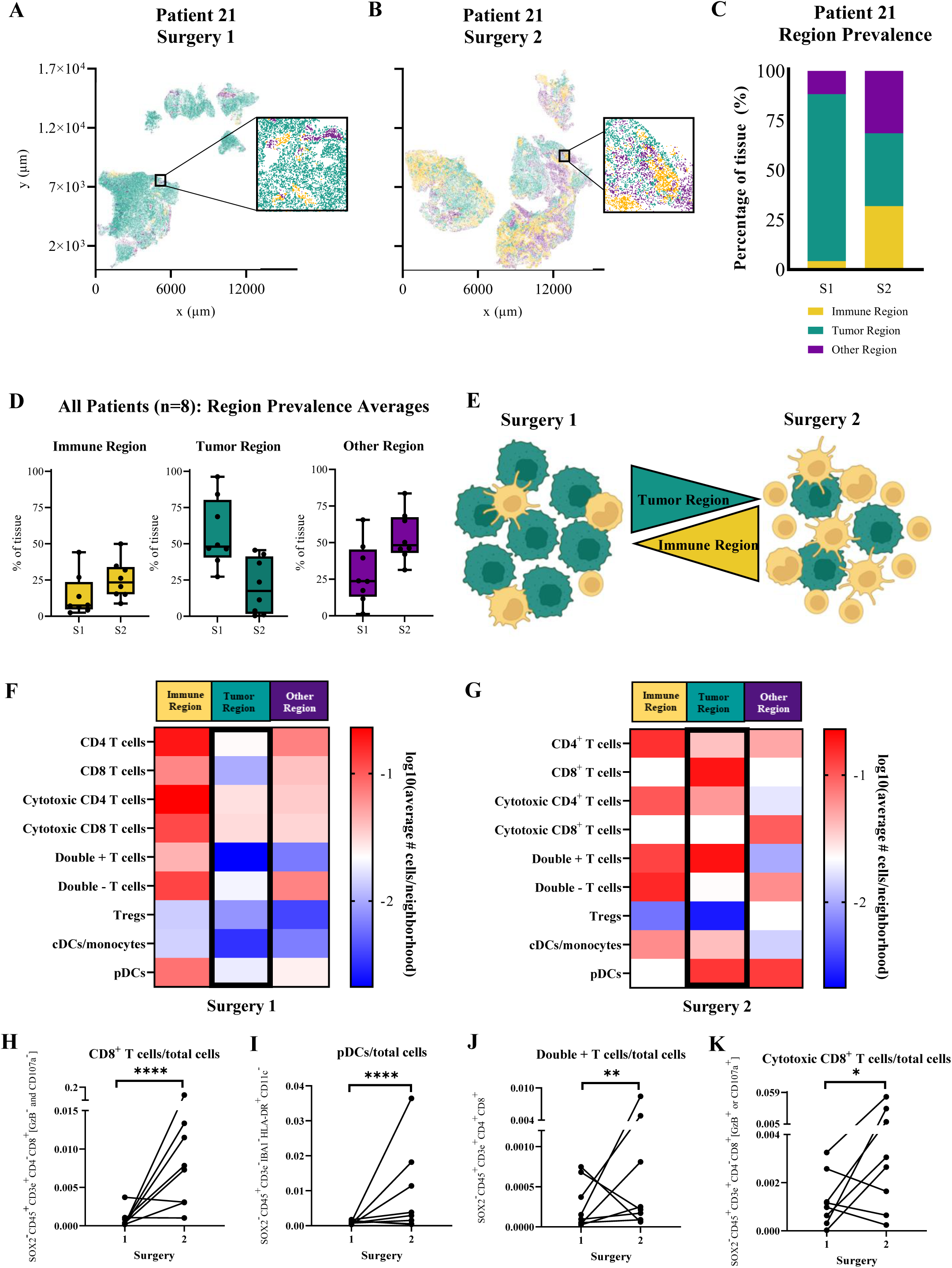
Neighborhood analysis of the tumor microenvironment in the biopsies obtained at primary tumor resection and post dual-vector TK-Flt3L treatment at tumor recurrence (see also Figure S9 and S10 of the Supplemental Appendix). The tumor microenvironment in HGG tissues was analyzed at primary tumor resection and at tumor recurrence using a neighborhood approach. Neighborhoods of 50μm were raster scanned and clustered into three distinct regions. The cells of primary tumor resection tissue for patient ID #21 are shown in a plot colored by their region type **(A)**, while cells of recurrent tumor resection tissue for the same patient are shown in a plot **(B),** (note: low magnification and multiple colors inter-mixing may create the appearance of more than three colors (Gold, Teal, and Purple) in the graphs; however there is no grey color in the image. The prevalence of the three region types within patient ID #21’s samples is illustrated in a bar graph **(C)**, Box plots displaying region prevalence in tissues across all patients. **(D)**. Cellular re-organization of immunologically cold (surgery 1) into hot and inflamed gliomas (surgery 2) post dual-vector TK-Flt3L treatment is illustrated in **(E)**. Heatmaps displaying cluster composition averaged across all primary tumor samples. Cluster composition is the average number of each cell type per neighborhood within a specific region **(F)** and all recurrent tumor samples **(G)** are also presented. Additionally, quantification of CD8+ T cells out of total cells (p < 0.0001) **(H)**, pDCs out of total cells (p < 0.0001) **(I)**, double positive T cells out of total cells (p=0.0022) **(J)**, and cytotoxic CD8+ T cells out of total cells (p=0.031) **(K)** is displayed in spaghetti plots for surgery 1 samples and surgery 2 samples. A generalized linear mixed effects regression model was employed to compare the rates of cell type counts between two different time points. The model assumed a negative binomial distribution, with a log link function and log (total cell) included as the offset in the regression. The results reported included the rate ratio (RR) and p-value. To control the family-wise type I error rate, the p-values were adjusted using the Sidak correction method.

More generally, further analysis revealed no differences between surgery 1 and 2, in cDCs/monocytes, double negative T cells, CD4^+^ T cells, cytotoxic CD4^+^ T cells, Regulatory T cells (Tregs), macrophages, or “other leukocytes” (CD45^+^ cells not expressing any other immune markers) (Figure S7B-H). However, the analysis showed statistically significant increases in CD8^+^ T cells (p<0.0001), pDCs (p<0.0001), double positive T cells (p=0.0022), and cytotoxic CD8^+^ T cells (p=0.031) when comparing data from surgery 1 to data from surgery 2, (post-radiotherapy, chemotherapy, and gene therapy) (Figure 4H-K). The co-localization of CD45 and HLA-DR expression was utilized to identify pDCs (Figure S8A), while the co-localization of CD45, CD3e, CD8, and CD4 expression was employed to identify double positive T cells (Figure S8B). CD8+ T cells were identified by co-localization of CD45, CD3e, and CD8; cytotoxic CD8+ T cells were identified by co-localization of these three markers, plus the additional co-localization of Granzyme B or CD107a (Figure S8C).

### Bioactivity of the viral gene therapy vectors: persistence of HSV1-TK immunoreactivity and increase in circulating level of Flt3L post dual-vector administration

We aimed to demonstrate the long-term expression of HSV1-TK and increase in Flt3L production in patients who received the dual-vector TK-Flt3L treatment through the analysis of biopsies and serum samples, respectively. In total, eight patients underwent two surgeries, and we found HSV1-TK positive cells in five of them through immunofluorescence and IHC (Patient IDs #2, 7, 8, 14, and 21). The longest period of expression was observed in patient #21, who exhibited expression for 17 months after treatment administration (Figure 5A and Figure S12). We validated the immunodetection’s specificity by performing pre-absorption of antibodies with Ad-TK infected cells; this eliminated the detection of HSV1-TK immunoreactivity as shown in Figure 5B and Figure S13. The specificity of the generated antibodies against HSV1-TK was evaluated using Western Blot analysis (Figure S15A of Supplemental Appendix). Further, the antibody’s specificity was tested in vivo and in vitro against infected and non-infected cells and tissues (Figure S15B and S15C of Supplemental Appendix). Further analysis of HSV1-TK immunoreactive cells at tumor recurrence showed three types of cells: tumor cells (cells that are positive for HSV1-TK+, SOX2+, and GFAP+), glial/astrocyte cells (cells that are HSV1-TK+, GFAP+ and SOX2-), and neurons, which were HSV1-TK+, GFAP- and SOX2-, and displayed typical neuronal morphologies of dendrites and axons (Figure 5C). We note that the continued presence of HSV1-TK expression in tumor cells supports the administration of Valacyclovir over longer time periods in future clinical trials.

**Figure 5:**
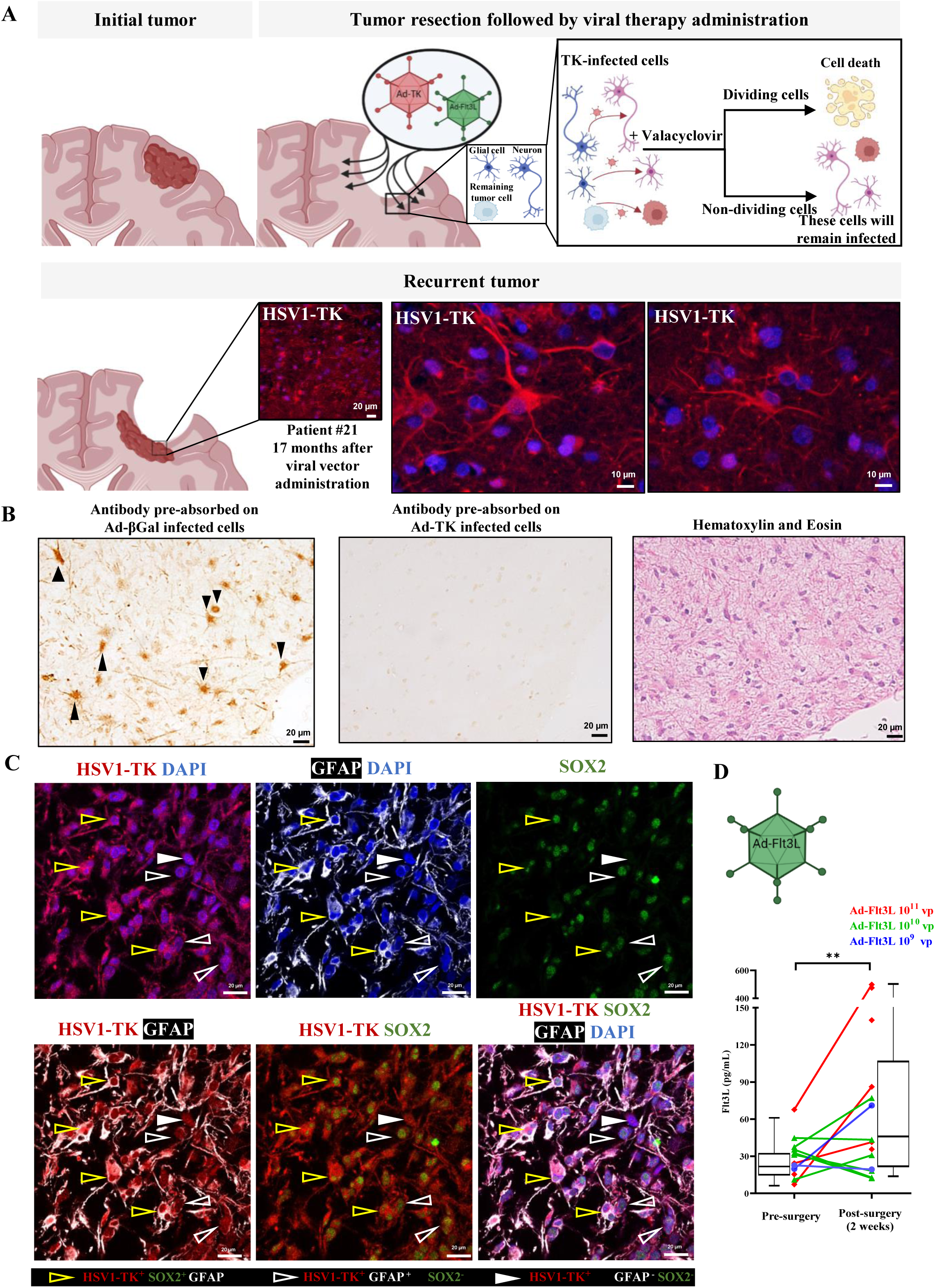
Persistent expression of HSV1-TK and Flt3L post-dual vector TK-Flt3L treatment at tumor recurrence (see also Figure S12 and S13 of the Supplemental Appendix). **A)** Top panel illustrates the administration of viral therapy in the cavity after the initial tumor resection, which infects not only the infiltrating tumor cells but also the surrounding brain parenchyma comprising glial cells and neurons. HSV1-TK infected cells that divide in the presence of Valacyclovir die; non-dividing cells such as neurons, or cells that did not divide during the Valacyclovir cycles will remain alive and infected. Bottom panel shows the infected cells that did not die during the dual vector TK-Flt3L treatment are still to be present at tumor recurrence, as evidenced by the positive immunoreactivity for HSV1-TK (red) in the immunofluorescence analysis of patient ID#21’s recurrent tumor biopsy 17 months following viral therapy administration. Five out of eight patients who had a second surgery showed a positive immunoreactivity for HSV1-TK (Figure S12A); sections from three negative patients are shown in Figure S12B. Nuclei are shown in blue (DAPI). **Panel B)** shows the same location of consecutive sections of surgery 2 of patient ID#21, where the specificity of the HSV1-TK antibody was tested by pre-absorbing it on (left section) infected cells with a control virus (βGal): Positive control; (middle section) infected cells with Ad-TK: Negative control, specific neutralization of the antibody while the right section shows the Hematoxylin and Eosin stain of the histology of the positive area (Lower magnification of the same area is shown in Figure S13). **Panel C**) presents the immunostaining of the tissue obtained at tumor recurrence for HSV1-TK (red), GFAP (white), and Sox2 (green) of patient ID#21. Nuclei are shown in blue (DAPI). Yellow arrows point to cells (presumptive tumor cells) that are positive for HSV1-TK+, SOX2+ and GFAP+, white unfilled arrows point at HSV1-TK+, GFAP+ and SOX2- infected astrocytes, white filled arrows point at HSV1-TK+, GFAP- and SOX2- (possibly neurons). **D)** shows the expression of circulating level of Flt3L in the patients at initial screening and 2 weeks after treatment (Patient IDs# 5, 7, 8, 10, 11, 12, 13, 14, 16, 18, 19, 20, and 21). The data analysis showed a significant increase in Flt3L levels after the dual vector TK-Flt3L treatment (**p=0.004). Statistical analysis was performed on R (version 4.2.1). Statistical significance was defined as p< 0.05. A linear mixed effects model was used to compare outcomes at the initial and post time points while considering the paired nature of the data measured at the two time points, and the outcome was log transformed to ensure goodness fit of the model.

To detect potential activity from the Ad-Flt3L vector, we measured the patients’ circulating level of Flt3L before (initial screening, pre-surgery) and two weeks after viral administration (Figure 5D) (Patient IDs# 5, 7, 8, 10, 11, 12, 13, 14, 16, 18, 19, 20, and 21). The results show that Flt3L circulating levels two weeks after treatment were statistically significantly higher than the initial value (p=0.004). Our data supports the hypothesis that the increase in Flt3L circulating levels in the patients is due to the expression from vectors injected into the brain.

### Anti-adenovirus antibodies

Neutralizing anti-adenovirus antibody titers were tested in cohorts B through F (n=15; Patient IDs# 5, 7, 8, 10, 11, 12, 13, 14, 16, 18, 19, 20, and 21) and the results summarized in Figure S14 of the Supplemental Appendix. The baseline anti-adenovirus antibody titers prior to dual viral vector injection were between <2 to 2048. Specifically, five patients (Patient IDs #11, 14, 16, 18, and 20) had no circulating anti adenovirus neutralizing antibodies (titer ≤ 2) at basal levels before delivery of vectors; four patients had titers between 8-64; and two patients had high basal levels (512-2048). All patients who were evaluated serially showed a significant increase in their antibody titer following the delivery of the viral vector (p=0.0005).

## Discussion

We report on the first-in-human phase 1 clinical trial of dual cytotoxic and immune-stimulatory adenoviral vector administration combined with SOC consisting of maximally safe resection followed by chemoradiation and adjuvant chemotherapy. The treatment was well tolerated, with no DLT attributable to the administration of adenoviral vectors. There were occasional mild to moderate toxicities that were possibly related to Valacyclovir. However, we cannot definitively conclude whether the toxicity of Valacyclovir was increased or decreased compared to the usual Valacyclovir toxicity in the absence of dual vector therapy, as our sample size (n=18) is insufficient to make a definite determination. Thus, reduced dose of Valacyclovir in future studies may be a consideration. An early study of adenovirus injected into the tumor cavity post-surgical excision found toxicity at 2×10^12^ pfu injected.^10^ Since this study, trials using adenoviral vectors, including ours, have remained one log below this dose, and consequently MTDs were not reached.

The tissue obtained at tumor recurrence (and post-dual vector TK-Flt3L treatment), showed an increase in CD8+ T cells as revealed by IHC except the decrease in CD8^+^ cells seen in patient ID #2 at recurrence. The finding was further confirmed by spatially resolved multiplex IHC, which showed an increase in CD8+ T cells expressing activation markers granzyme B and CD107a. This might suggest a differential response to the therapy in Anaplastic Ependymoma compared to HGG. Future investigations and studies will be required to explore the underlying mechanisms and implications of this specific response pattern. Additionally, there was a significant increase in double positive T cells, which could be pro- or anti tumor.^43^ An increase of pDCs was also observed, which was consistent with our preclinical findings.^15, 44^ Notably, there were no significant differences found in macrophages or Tregs between the primary tumor and tumor recurrence when all eight patients were considered together. However, five out of eight patients demonstrated reduced proportions of Tregs in their recurrent TME, as depicted in Supplemental Appendix (Figure S7G). Though these results do not agree with previous findings on the TME of GBM at recurrence (which demonstrated a substantial decrease in glioma-associated macrophages^45^ and an increase in Tregs^46^) they may be a result of our gene therapy, and thus indicate the induction of a hot TME. While some studies have reported an increase in CD8+ T cells at recurrence in response to SOC,^47, 48^ the magnitude of increase seen in our trial is numerically higher. For example, Mohme et al.^47^ looked at differences in CD8^+^/CD3^+^ ratios between primary and recurrent GBM. They found a 5% average increase in CD8^+^ cells within the CD3^+^ compartment. When we calculate these ratios with our data, we find a 28% increase in CD8^+^ cells within the CD3^+^ compartment. By quantifying the subsets of immune cells and conducting a cell neighborhood analysis, we demonstrate that immune infiltration occurs both around and within regions densely populated with tumor cells. Additionally, Treg and macrophage populations in the tissues examined did not increase, which differs from literature data indicating that they increase at recurrence. Thus, our data appear to be qualitatively different from previous assessments, a difference that could be due to the administration of our viral vectors. However, macrophages could still become immunosuppressive, and thus inhibit the anti-glioma immune response. Our cell-cell correlation analysis demonstrates an increased interaction of macrophages with T cells at surgery 2 corresponding with decreased interactions between macrophages and tumor cells (Supplemental Appendix, Figure S11), potentially suggesting that immunosuppressive macrophages could reduce the therapeutic impact of the increased number of CD8+ T cells and pDCs, as recently described.^49, 50^

Although this study was not powered for survival analysis, the median OS of 21.3 months (95%CI: 11.1, 26.1) is promising, especially considering that GBM has a known historical OS rate of 14-16 months, though high complexity centers report survivals of 19-21 months.^51^ Remarkably, the two-year survival rate was 38.8% (7/18 patients), the three-year survival rate 16.66% (3/18 patients), and the four-year survival rate was 11.11% (2/18 patients); one patient is still alive 57 months after vector administration (patient ID #16). The range of survival values at 2 years in four previous trials using Ad-TK+prodrug was 33-42.86%, and the range of overall survival (OS) was 10.66-25.3 months.^9–11, 38, 52, 53^ Future directions may consider the inclusion of immune checkpoint blockade^54^ and/or methods to reduce the strong immunosuppressive glioma microenvironment.

Since the modern dawn of gene therapy in the 70’s there have been close to sixty viral gene therapy trials for brain tumors using adenoviral vectors (∼20 trials), HSV1 vectors (∼12 trials), retroviral vectors (∼6 trials), and measles virus, Newcastle Disease virus, vaccinia, reovirus, Parvovirus-H1, and Poliovirus derived vectors.^8, 55^ Viral vectors have been used to deliver conditional cytotoxicity, oncolysis, cytokines, and the use of checkpoint inhibitors in combination with various viral vectors.^8, 55^ In gene therapy trials, clinical outcomes have been less impressive than expected, with only small numbers of patients displaying long-term survival. A recent publication utilizing up to six repeated injections of intratumoral oncolytic HSV-G47Δ injections for the treatment of residual or recurrent glioblastoma in a phase 2 trial reported promising one year survival rates.^56^

The results of a phase 1 trial by Chiocca et al.^57^ demonstrated the feasibility and tolerability of a single agent injected into the resection cavity walls. In this study, the investigators delivered an adenoviral vector expressing a regulated form of IL-12 in patients with recurrent HGG. In a subsequent phase 1 trial, they explored the combination of this treatment with Nivolumab, an immune checkpoint inhibitor.^58^ The combination approach showed promising antitumor immune responses, regulated by the oral administration of Veledimex, which controlled IL-12 transcription. While the trials reported some degree of moderate toxicity, they highlighted the importance of tightly controlling the release of potent proinflammatory cytokines such as IL-12.^57, 58^ Furthermore, PsP has been reported in various trials using Oncolytic virus or viral vectors.^39, 56, 59–61^ While the incidence of PsP appears lower in our trial, a proper confirmation will need to await larger phase 2 or 3 comparative trials.

Correlative studies have yielded more promising results, with several indicating an increase in immune cells within areas of the brain/tumor that had previously been treated with gene therapy vectors, potentially transforming “cold” tumors into “hot” ones.^62, 63^ Our analysis of cellular neighborhoods and spatial phenotyping also revealed a significant increase in immune cell infiltration during the second surgery as compared to pre-treatment biopsies. In fact, five out of the eight consecutive biopsies showed immune infiltration, successfully converting “cold” GBM to “hot” and inflamed, as illustrated in Figure 4E and Supplemental Appendix (Figure S9-S11).

We detected an increase in circulating antibody titers following the delivery of the viral vector (p=0.0005). This indicates that following peri-tumoral injection of adenoviral vectors it is possible for the humoral immune response to increase. There was no correlation between vector dose, serum antibody levels, OS, or intracranial quantitative immune responses measured by IHC. Interestingly, HSV1-TK immune reactivity was detected in a patient who seroconverted (Patient ID #14). Thus, seroconversion did not abolish our capacity to detect the presence of the HSV1-TK transgene.

Our approach to GBM treatment utilizing gene therapy exhibits several novel and distinctive characteristics. Our gene therapy trial employs two distinct adenoviral vectors: one expressing HSV1-TK, and the other expressing Flt3L. As illustrated in Figure 1, the most original aspect of our approach involves the use of Flt3L to attract and differentiate DCs within the tumor’s immune microenvironment (TiME). Interestingly, the combination of HSV1-TK plus Valacyclovir kills tumor cells, thereby releasing tumor antigens and DAMPs. Consequently, these antigens and DAMPs become available to DCs, which take up tumor antigens, undergo activation by DAMPs, and subsequently migrate to lymph nodes to stimulate antigen-specific T cells. All of these mechanisms have been previously described by us in preclinical animal models of glioma.^13, 15, 22–26, 31, 62, 64–68^

Moreover, we extended the administration of the HSV1-TK substrate, Valacyclovir, to two cycles. The rationale for extending the administration of Valacyclovir is based on preclinical data demonstrating that in rodents, adenoviral vector mediated transgene expression can last up to one year.^35–37^ As the cytotoxic effect is only realized during the administration of Valacyclovir and tumor cell division, it might be sensible to further extend Valacyclovir administration to increase the cytotoxic potential of the HSV1-TK/Valacyclovir combination. Additionally, it could be equally important to consider multiple vector administrations, as recently described.^56^

Remarkably, tumor tissue obtained at tumor recurrence showed persistent expression of HSV1-TK up to 17 months post dual-vector administration. Five out of eight patients displayed HSV1-TK immunoreactive cells in tissue obtained from surgeries on tumor recurrences, and increased levels of circulating Flt3L were observed two weeks after treatment. These findings support the use of extended administration of Valacyclovir in a phase 1b/2 clinical trial. The sustained expression of HSV1-TK plus future prolonged administration of Valacyclovir may be a mechanism for maintaining disease control over a prolonged period, as it stimulates systemic antitumor immunity. The continued increase in the amount of T cells and DCs at second surgeries suggests that such an immune response might be at work. Both the longevity of transgene expression, and the increase of immune cells, corroborates our preclinical data.^13, 15, 22, 24, 27, 31^ The promising survival data and recruitment of immune cells, along with the persistent transgene expression from the vectors, suggest that this treatment approach may have significant potential for the management of the disease and warrants further investigation in a phase 1b/2 clinical trial.

## Data Availability

All data produced in the present work are contained in the manuscript and are also available upon reasonable request to the authors.

## Ethics Statement

We have obtained ethics approval from the Institutional Review Board-MED at the University of Michigan School of Medicine before enrollment (HUM00057130). The trial has been registered at ClinicalTrials.gov (NCT01811992). Prior to the initiation of the study, we obtained an Investigational New Drug (IND) from the FDA (BB-IND 14574). All patients provided pre-operative consent, and definitive enrollment occurred intraoperatively after confirming the presence of malignant glioma through Pathology examination. The most recent version of the Clinical Protocol can be accessed in the study protocol document.

## Authors’ contributions

Conceptualization and Study Design: MGC and PRL. Patients’ enrollment and treatment: YU, DO, LJ, JH, OS, DL, AM, SHJ, WA, TH, KS, and KM. Neuropathology: SCP, APL, PM, and SV. Radiotherapy: MMK, DRW, and TL. Exploratory immunological and histopathological analysis: MLV, MEJW, SMF, AC, DZ, VNY, PD, MGC, and PRL. Figures: YU, MLV, MEJW, SMF, AC, MGC, and PRL. Data curation: YU, MLV, MEJW, SMF, AC, DA, RK, MGC, and PRL. Statistical analysis: YU, and LZ. Analysis and data interpretation: YU, LJ, AM, MLV, MEJW, SMF, AC, LZ, MGC, and PRL. Writing-original draft: YU, SMF, MGC, and PRL. Writing- review and editing: All authors thoroughly reviewed the data, made valuable contributions to the development of the manuscript, and provided their approval for the final version to be published. Data Validation: YU, MLV, MEJW, SMF, AC, LZ, MGC, and PRL. Fund Raising: MGC and PRL.

## Acknowledgements

Funded in part by Phase One Foundation, Los Angeles, CA, The Board of Governors at Cedars-Sinai Medical Center, and The Rogel Cancer Center at The University of Michigan.

[clinicaltrials.gov: NCT01811992]. We express our sincere gratitude to the patients and their families for their invaluable participation in this trial.

## SUPPLEMENTAL APPENDIX

### Supplementary Methods: Eligibility

#### Inclusion Criteria

1. Newly identified supratentorial brain lesion compatible with a diagnosis of high-grade glioma (WHO III or IV) by MR with no prior treatment with either gene therapy, chemotherapy or radiation treatments that is amenable to attempted gross total resection (GTR). “High grade glioma” can include glioblastoma multiforme (WHO grade IV); gliosarcoma (WHO grade IV); anaplastic astrocytoma (WHO grade III); anaplastic oligodendroglioma (WHO grade III); and anaplastic ependymoma (WHO grade III).
2. Intraoperative histological frozen section analysis at the time of tumor resection shown to be compatible with high-grade glioma or malignant glioma (as described elsewhere throughout this document). If intraoperative diagnosis was not compatible with high grade glioma, the patient was not enrolled.
3. Karnofsky Performance Status (KPS) score ≥70
4. Laboratory values
5. CBC/differential obtained within 14 days prior, with adequate bone marrow function were defined as follows:
  1. Absolute neutrophil count (ANC) ≥ 1,500 cells/mm3; Platelets ≥ 100,000 cells/mm3; Hemoglobin ≥ 10.0 g/dl (Note: The use of transfusion or other intervention to achieve Hgb ≥10.0 g/dl is acceptable.);
  2. Adequate renal function, as defined below: BUN ≤ 30 mg/dl within 14 days prior. Creatinine ≤ 1.7 mg/dl within 14 days prior.
  3. Adequate hepatic function, as defined below: Bilirubin ≤ 2.0 mg/dl within 14 days prior. ALT/AST ≤ 3x laboratory upper limit of normal within 14 days prior.
6. Male and female; both genders must use contraception if of reproductive capacity
7. For women of childbearing potential, a negative pregnancy test performed within 14 days of surgery
8. Capable of informed consent
9. 18-75 years of age

#### Exclusion Criteria

1. Diffusely multifocal lesion
2. Lesions not amenable to GTR
3. Tumors infiltrating the cerebellum, bilateral corpus callosum (“butterfly glioma”), ventricular system, or brain stem
4. Infratentorial high grade glioma
5. Primary central nervous system (CNS) disease that would interfere with subject evaluation
6. Prior history of other cancer within five years except curative cervical cancer in situ, basal or squamous cell carcinoma of the skin
7. Evidence of other significant disease including hematologic, renal or liver disease that is not explained by the patient’s current medical condition or concomitant disease
8. Patient known to have HIV, Hepatitis B, Hepatitis C
9. Active systemic infection
10. Immunosuppressive disorders (i.e., chronic steroid therapy, acquired or congenital immune deficiency syndromes, autoimmune disease)
11. Serious medical conditions (i.e., CHF, angina, diabetes mellitus Type 1, COPD, bleeding diathesis)
12. Any contraindication for undergoing MRI
13. Pregnant or lactating females
14. Unacceptable anesthesia risk
15. Evidence of bleeding diathesis or use of anticoagulant medication or any medication that may increase the risk of bleeding that cannot be stopped prior to surgery
16. Prior gene therapy
17. Allergy to Valacyclovir
18. Unable to take oral tablets

### Supplementary Methods: Schedule of Assessments

|  | Screening (<14 days prior to surgery) | Day 0 (prior to surgery) | Day 0 (during surgery) | Day 2 ± 1 day | Week 2 ± 1 wk Neurosurgery visit | Week 1-3 NeuroOnc visit | Week 6 ± 2 wk | Week 10 ± 2 wk | Week 14 ± 2 wk | Week 18 ± 2 wk | 12-Months Follow up<sup>8</sup> | 24-Months Follow up<sup>8</sup> | Overall Survival<sup>8</sup> |
| --- | --- | --- | --- | --- | --- | --- | --- | --- | --- | --- | --- | --- | --- |
| Medical history | X |  |  |  | X | X | X | X | X | X |  |  |  |
| Physical exam | X |  |  |  | X | X | X | X | X | X |  |  |  |
| Karnofsky Performance Status | X |  |  |  | X | X | X | X | X | X |  |  |  |
| Vital signs<sup>1</sup> | X | X |  |  |  | X |  |  |  |  |  |  |  |
| Hematology<sup>2</sup> | X |  |  | X | X |  |  |  |  |  |  |  |  |
| Clinical chemistry<sup>3</sup> | X |  |  | X | X |  |  |  |  |  |  |  |  |
| MRI | X |  |  | X<sup>5</sup> |  |  | X | X | X | X |  |  |  |
| ECG | X |  |  |  |  |  |  |  |  |  |  |  |  |
| Pregnancy test (if applicable) | X | X |  |  |  | X | X | X | X | X |  |  |  |
| Anti-Ad Abs | X |  |  |  | X |  | X | X |  |  |  |  |  |
| Serum sample (stored) |  |  |  |  | X |  | X | X |  |  |  |  |  |
| Blood sample (PMBC) | X |  |  |  | X |  | X | X |  |  |  |  |  |
| Neurological assessments<sup>7</sup> | X | X |  | X | X | X | X | X | X | X |  |  |  |
| Histological confirmation of high-grade glioma |  |  | X<sup>4</sup> |  |  |  |  |  |  |  |  |  |  |
| Vector delivery |  |  | X |  |  |  |  |  |  |  |  |  |  |
| Start Valacyclovir<sup>6</sup> |  |  |  | X Round 1 |  |  |  | X Round 2 |  |  |  |  |  |
| Concomitant medications |  | X |  | X | X | X | X<sup>8</sup> | X | X<sup>8</sup> | X |  |  |  |
| Adverse events |  |  |  | X | X | X | X<sup>8</sup> | X | X<sup>8</sup> | X |  |  |  |
| Survival follow up |  |  |  |  |  |  |  |  |  |  | X | X | X |

1. Vital signs include body weight, temperature, blood pressure, heart rate, respiration rate. 2. Hematology panel to include CBC, WBC, Differential, PT/PTT (PT/PTT only at screening). 3. Clinical Chemistry includes glucose, calcium, albumin, total protein, electrolytes [sodium, potassium CO2, chloride], BUN, creatinine, alkaline phosphatase, ALT/SGPT, AST/SGOT, bilirubin. 4. Confirmation of malignant glioma leads to formal patient enrollment, and triggers preparation of viral vectors for treatment administration. At this time, surgery informs O-CTSUS that biopsy confirms histological eligibility, and treatment proceeds. 5. Post-op MRI to determine the degree of tumor resection was obtained within the first 48 hours postoperatively. 6. Valacyclovir was administered orally at 2g, 3 times daily for 14 days beginning 1-3 days after vector injection per clinician judgement. Round 2 entailed 14 days of treatment, 2g 3x/day. 7. Neurological Assessments include Mini Mental Status. 8. Survival follow-up information could be assessed by telephone calls, patient medical records, and/or clinic visits. If the patient withdrew from study, the study staff would have used a public information source (e.g., county records) when permissible, to obtain information about survival status only.

### Supplementary Methods: Protocol Treatment Plan

#### Administration of Ad-hCMV-TK and Ad-hCMV-Flt3L

The total volume of Ad-hCMV-TK and Ad-hCMV-Flt3L injected was 2 ml.

#### Surgical Procedure

A craniotomy with resection of the tumor was performed. At the neurosurgeon’s discretion, this involved stereotactic methods and/or intraoperative navigational guidance and/or intraoperative MRI or other radiologic guidance. After tumor resection had been completed and adequate hemostasis obtained, two tuberculin syringes were used to deliver the Ad-hCMV-TK and Ad-hCMV-Flt3L vector in 100 microliters at each of twenty sites. The assignment of dosages was determined preoperatively as per protocol (Table S1). Injections were done over 3-5 minutes for each syringe and each vector was delivered at random sites throughout the cavity. The vectors were brought from University of Michigan Research Pharmacy in sterile syringes as prepared under the supervision of a Research Pharmacist. Syringes containing vectors were delivered on ice to the OR. The surgeon placed a sterile 25-gauge needle on the syringe within the OR. The syringes did not remain in the OR for more than 4 hours before use. The injection sites were selected by the surgeon to avoid injections into adjacent motor or speech cortex, the cerebral ventricle, or seepage into the subarachnoid space. A total of 2ml of vectors will be injected as 0.1 ml/injection site across 20 sites. No injections were made into mapping-defined areas of eloquent brain to avoid any risk of injection-related complications in these areas. In addition, surgeons maintained a safety margin, such that injections of vectors were made at least one gyrus away from regions of eloquent brain. After the injections were complete, the needle was slowly withdrawn, and the remainder of the operation consisted of routine wound closure. The delivery of the viral vectors required an additional 10 minutes to the time of surgery.

#### Valacyclovir Administration

Valacyclovir treatment began 1-3 days after vector administration, at a dose of 2 grams given orally three times per day for a course of 14 days. A second course of Valacyclovir was given beginning week 10 (+/1-2 weeks). Valacyclovir was dispensed from the University of Michigan Research Pharmacy with a medication diary, which was turned in by the patient to the study team at the end of each course of Valacyclovir.

The capacity of Ad-hCMV-TK to kill glioma cells depends on three factors that need to be present simultaneously:

- the glioma cells need to express HSV1-TK;
- the nucleoside analogue Valacyclovir needs to be present; and
- the cell must enter cell division (mitosis).

The second cycle of Valacyclovir is designed to target and kill residual tumor cells infected with Ad-hCMV-TK that were not dividing during the first cycle. As the population of cells that are expressing TK that do not divide early is likely to be significant (based on patterns of glioma recurrence), the second cycle of Valacyclovir may be able to kill these cells, and thus increase the potential to present antigens and toll receptor ligands to immune cells recruited to the tumor microenvironment by Ad-hCMV-Flt3L. The dose/schedule of Ad-hCMV-TK has been used in clinical trials for prostate, ovarian, and malignant glioblastoma. The dose of Ad-hCMV-Flt3L was determined in this clinical trial.

#### Radiation Therapy (standard of care)

Radiotherapy (RT) was determined and administered as per standard-of-care for the individual patient by the treating radiation oncologist. For most patients, RT began 15-35 days after Ad-hCMV-TK and Ad-hCMVFlt3L injection. It consisted of standard external field radiation, limited to the area of the tumor and brain adjacent to the tumor. The most common regimen was 60 Gy, delivered as 2 Gy per day, five days/week for six weeks. Another standard regimen was 40.05 Gy, delivered as 2.67 Gy per day, five days/week for three weeks. Neither the performance of radiation, the timing, nor the details was directed by this protocol.

#### Temozolomide Therapy (standard of care)

Temozolomide capsules are an approved oral chemotherapeutic drug for the treatment of adult patients with newly diagnosed high grade glioma concomitantly with radiotherapy and then as maintenance treatment. Treatment with temozolomide was determined as per standard-of-care for the individual patient by the treating neuro-oncologist. It was anticipated that, for most patients, concomitant temozolomide would be taken with RT starting 15-35 days after Ad-hCMV-TK and Ad-hCMVFlt3L injection. It was also anticipated that, for most patients, the duration of concomitant temozolomide would coincide with the duration of RT, as described above. It was also anticipated that neither the performance of radiation, the timing, nor the details were directed by this protocol. Pneumocystis carinii (PCP, also known as Pneumocystis jirovecii) Prophylaxis. Some physicians recommend trimethoprim/sulfamethoxazole or other medications as prophylaxis for PCP for patients treated with temozolomide. There is no national standard-of care and no standard practice at the University of Michigan. The use of PCP prophylaxis was not directed by this protocol.

#### Maintenance Phase (Standard of Care)

Standard-of-care treatment generally includes cyclic temozolomide, starting about 4 weeks after completion of RT with concomitant temozolomide, take on five successive days every 28 days (1 cycle) for 6-12 cycles, if the tumor has not progressed. It was anticipated that most patients in this protocol would take 1-2 cycles of temozolomide during the 16-week period of intense monitoring. Decisions about cyclic temozolomide were made by the treating neuro-oncologist per standard of care. Neither the administration of cyclic temozolomide, its timing, nor its details are directed by this protocol.

### Supplementary Methods: Toxicities and Dose Delays

#### Dose -Delays/ Potential Complications Specific to Adenoviral Vector Infusion

Fever may occur due to the local inflammatory and immune response to Ad-hCMV-TK and AdhCMV-Flt3L; acetaminophen will be used as necessary. If grade 2 or greater NCI CTCAE v.4 criteria for infection of the surgical wound is present (i.e., requiring systemic antibiotics therapy for treatment), Valacyclovir was not started or stopped. If the course of Valacyclovir had not yet been started, then the patient would receive the full dose. If the course of Valacyclovir had been stopped, then the patient would receive the remainder. Any such dose modification was noted.

In previous clinical trials evaluating Ad-hCMV-TK followed by prodrug administration, toxicity had been rare and the overall approach had been well tolerated. In a similar study in recurrent brain tumors, CNS toxicity (lethargy, confusion, grade 3 hyponatremia, fever, leukocytosis, intratumoral hemorrhage, and hydrocephalus) was observed in the 2×10^12^ vp dose level. In the lower dose levels, no DLTs were observed; however transient and less severe events included seizure, small intratumoral hemorrhage, worsening of neurologic symptoms, mild hyponatremia. However, as the combination of Ad-hCMV-TK and Ad-hCMV-Flt3L in combination with surgery, RT, and chemotherapy had not yet been evaluated in human clinical trials, we chose to evaluate in the Phase 1 study dose levels of Ad-hCMV-TK that had been shown to be well tolerated over a number of studies.

#### Valacyclovir

The most common adverse events with Valacyclovir reported in the prescribing literature are nausea in 6-15% of patients, headache in 14-35%, vomiting in 1-6%, dizziness or abdominal pain in 2-11%, arthralgia in 1-6% and depression in 1-7%. Laboratory abnormalities such as abnormal liver function tests had been observed in 1-4% of patients, while anemia, leucopenia, thrombocytopenia and elevated serum creatinine occurred in 1% of patients or less. In addition, the following adverse events have been identified in clinical practice: allergic reactions (anaphylaxis, angioedema, dyspnea, pruritus, rash and uticaria), CNS symptoms (confusion, insomnia, hallucinations, agitation, aggressive behavior, mania), gastrointestinal (diarrhea), renal failure, erythema multiforme, facial edema, hypertension and tachycardia. A rare but serious adverse event of thrombotic thrombocytopenia purpura/hemolytic uremic syndrome (TTP/HUS), in some cases resulting in death, has been observed in clinical trials of high dose Valacyclovir (8 grams per day) in patients with advanced HIV infection, and in allogeneic bone marrow transplant and renal transplant recipients.

#### Valacyclovir Dose Modifications (Dose Hold, Delay or Discontinuation) due to AEs

Subjects who experience a Grade 3 or greater NCI CTCAE v.4 Thrombotic thrombocytopenia purpura or Grade 2 or greater Hemolytic uremic syndrome or Grade 3 or greater serum creatinine were managed as outlined below in Tables S2 and S3. Adjustments to the dose reduction guidelines was based on the clinical judgment of the Investigator with approval by the Medical Monitor.

- For all patients starting Valacyclovir, baseline creatinine within 3 d before start of treatment, were measured.
- For patients who started with a normal/baseline creatinine level, recheck at day 7 + 2 d, was performed.
- For patients who started with elevated creatinine levels, recheck at least twice a week (every 4 d) through the 14 days, was performed.

If dosing was delayed for four weeks or longer, the cycle of Valacyclovir was discontinued.

For patients starting cycle 2 of Valacyclovir who had creatinine elevation during cycle 1, the following criteria were used:

- If max creatinine was Grade 1, start at dose level 0 or lower at discretion of investigator; if started lower and tolerated, increase one level at discretion of investigator.
- If max creatinine was Grade 2, start at dose level 1 or lower at discretion of investigator; if tolerated, increase one level at discretion of investigator.

#### Temozolomide

Temozolomide is a standard-of-care treatment for glioblastoma. Its use was not mandated or directed by the protocol, but it was expected that most patients would receive it. In patients receiving temozolomide, toxicities known compatible with temozolomide as the cause will not be considered as DLTs from the treatments administered as part of this study.

Patients treated with temozolomide have reportedly experienced myelosuppression including prolonged pancytopenia, which may result in aplastic anemia, which in some cases has resulted in a fatal outcome. In some cases, exposure to concomitant medications associated with aplastic anemia including carbamazepine, phenytoin, and sulfamethoxazole/trimethoprim complicates assessment. Geriatric patients and women have been shown in clinical trials to have a higher risk of developing myelosuppression.

Cases of myelodysplastic syndrome and secondary malignancies, including myeloid leukemia, have also been observed. Prophylaxis against *Pneumocystis carinii* pneumonia will be left to the discretion of the treating physician. There may be a higher occurrence of PCP when temozolomide is administered during a longer dosing regimen. However, all patients receiving temozolomide, particularly patients receiving steroids, should be observed closely for the development of PCP regardless of the regimen.

Grade 3/4 neutropenia occurred in 8% and Grade 3/4 thrombocytopenia in 14% of patients treated with temozolomide. The most common side effects associated with temozolomide therapy are nausea, vomiting, anorexia, alopecia, headache, fatigue, constipation, convulsions, weakness, and thrombocytopenia.

### Supplementary Methods: Clinical and Imaging Evaluation

Subjects were followed closely with clinical and radiographical assessments for 30 days after the completion of the second Valacyclovir course, approximately 16 weeks from the dual vector administration, and thereafter for survival. Disease progression was determined using the immunotherapy response assessment in neuro-oncology (iRANO) criteria.^1^ Detailed Schedule of Assessments can be found in the Supplemental Methods Section in the Supplementary Appendix.

The relationship between pre-operative diffusion-weighted imaging (DWI) and dynamic susceptibility contrast (DSC) perfusion-weighted imaging parameters and survival were analyzed using Pearson’s correlation analyses. The mean apparent diffusion coefficient (ADCmean), relative cerebral blood flow (rCBF), and relative cerebral volume (rCBV) were measured using volume-of-interest placed on the enhanced areas of the tumors. Normalized ADCmean (nADCmean), normalized rCBF (nrCBF), and normalized rCBV (nrCBV) were calculated by dividing by the reference values which were measured using three separate regions-of-interest placed in the normal-appearing white matter.

### Supplementary Methods: Production of anti-HSV1-TK antibody

A rabbit polyclonal antibody specific to HSV1-TK was generated in-house, by BioSource International Inc., CA using standard methods. Serum samples were collected after immunization. The specificity of the generated antibodies against HSV1-TK was evaluated using Western Blot analysis (Figure S15A of Supplemental Appendix). Further, the antibody’s specificity was tested *in vivo* and *in vitro* against infected and non-infected cells and tissues (Figure S15B and S15C of Supplemental Appendix).

### Supplementary Methods: Correlative Studies

#### Multiplex immunofluorescence and analysis using the Akoya PhenoCycler-Fusion

Multiplex immunofluorescence of all FFPE tissue (Figure 4) was performed on a PhenoCycler-Fusion by Akoya Biosciences per manufacturer’s instructions with the following addition. After antigen retrieval, a photobleaching protocol was performed to reduce autofluorescence. Briefly, slides were submerged in a solution of 30% w/v hydrogen peroxide, 1x PBS and 1M sodium hydroxide and sandwiched between two full spectrum LED light boxes for thirty minutes. Samples were then equilibrated in four three-minute 1x PBS washes before continuing with the remainder of the Akoya protocol. All the antibodies, except for the Anti-Hu IBA1 antibody, were obtained from Akoya Biosciences, Inc. The Anti-Hu IBA1 antibody was a custom conjugate created using an Abcam antibody (catalog number Ab221790) and was visualized with an Alexa Fluor™ 488 reporter. Further details regarding the antibodies used can be found in the supplemental appendix (Table S7). For the Akoya protocol, samples are stained with antibodies conjugated to unique oligonucleotide sequences (i.e., barcodes). Reporters, consisting of fluorophores attached to oligonucleotide sequences complementary to those on the barcodes, are hybridized and imaged three at a time in consecutive cycles on the Phenocycler-Fusion platform.

After completing the PhenoCycler-Fusion run, we imported the QPTIFF images into QuPath version 0.4.0 for sample annotation, cell segmentation, and phenotyping.^2^ Initially, we included all tissue from the image in the annotation. We then excluded areas with artifacts and putative non-tumor tissue. Exclusion was determined based on marker expression (SOX2, GFAP, and vimentin) and cell density. After annotation, segmentation was performed on the final annotation using the StarDist segmentation script.^3^ We carried out phenotyping using a hierarchical scheme implemented with a script using single intensity thresholds for each marker as shown in Supplemental Appendix (Figure S7A). The phenotyping was performed on approximately 4.1 million total cells from 16 samples.

Once the classification of all cells was complete, we exported the data into CytoMAP for neighborhood analysis. We defined raster scanned neighborhoods with a 50-micron radius and clustered resulting neighborhoods into regions. The Davies Bouldin criteria was used to determine the number of regions, and the NN self-organizing map algorithm was employed for clustering the neighborhoods into regions. We then analyzed region statistics to investigate changes in neighborhood composition using cluster composition. In CytoMAP, cluster composition is defined as the average number of a particular cell type in neighborhoods within a given region. For instance, we analyzed the average number of CD8 T cells per neighborhood within a specific region.

#### Statistical Analysis

A generalized linear mixed effects regression model was employed to compare the rates of cell type counts between two different time points. The model assumed a negative binomial distribution, with a log link function and log (total cell) included as the offset in the regression. The results reported included the rate ratio (RR) and p-value. To control the family-wise type I error rate, the p-values were adjusted using the Sidak correction method.

#### Statistical analysis of immunohistochemistry

For each marker (i.e., CD8), a linear mixed effects (LME) model was used to compare NumPos (log-transformed) between two surgery procedures. Random effect was included in the LME model to consider correlations between data measured on the same subject. Significance is determined if p<0.05. All analyses were conducted using SAS (version 9.4, SAS Institute, Cary, NC).

#### Antibody measurements

Prior to viral vector injection, circulating neutralizing anti-adenovirus antibody titers were measured,^4–6^ then at week 1, 2, 6, 10 and 48. The titers were expressed as follows: 1:16, is expressed as 16, etc.; the higher the number the higher the anti-adenoviral titer. The neutralizing anti-adenovirus antibody titers were measured in patients’ serum samples (Patient IDs# 5, 7, 8, 10, 11, 12, 13, 14, 16, 18, 19, 20, and 21). Sera were heat-inactivated at 56 °C for 30 min and serially diluted 2-fold in DMEM with 10% FBS 12 times (1:2- 1:4096). Each dilution was incubated with 1×10^6^ i.u. of RAd35 (Ad-hCMV-LacZ) for 90 min at 37 °C. Volumes of 50 µL of each dilution were transferred to 96-well plate containing 4 x 10^4^ HEK 293 cells for 60 min at 37 °C (performed by triplicates), followed by a 50 µL addition of DMEM with 10% FBS in each well. After 20h at 37 °C cells were fixed with 4% paraformaldehyde in PBS and stained with 5-bromo- 4chloro-indolyl-ß-D-galactoside (X-gal). The neutralizing anti-adenovirus antibody titers for each sample was defined as the reciprocal of the highest dilution of serum at which 50% of RAd35-mediated transduction was inhibited.

### Supplementary Results: MRI Studies

An association between lower ADC (apparent diffusion coefficient) values and poor survival have been reported in glioblastoma.^7–9^ A systematic review and meta-analysis data also suggest significant correlation of pretreatment low ADC values with poor survival in patients with recurrent HGG treated with bevacizumab.^9^ Given the heterogeneity of the treatment course among the patients in this study such as additional resection that modify the appearance of target lesion at the follow up imaging, the pre-operative baseline DWI and DSC parameters were analyzed. Although no significant correlations were found between MRI parameters and survival, there were weak correlations between baseline pre-operative nrCBV and OS, and between pre-operative nADC mean and OS.

**Table S1.** Dual Viral Vector Dosing.

| Cohort | Ad-hCMV-TK<br>(10<sup>N</sup> vp) | Ad-hCMV-Flt3L<br>(10<sup>N</sup> vp) |
| --- | --- | --- |
| A | 10 | 9 |
| B | 11 | 9 |
| C | 10 | 10 |
| D | 11 | 10 |
| E | 10 | 11 |
| F | 11 | 11 |

**Table S2.** Dose Modification for Valacyclovir.

| Dose level |  |
| --- | --- |
| 0 (starting) | 2 g TID |
| -1 | 2 g BID or 1 g TID |
| -2 | 1 g BID or 0.5 g TID |

**Table S3.** Renal Adverse Event Management Algorithm for Valacyclovir.

| Grade of Creatinine Elevation<br>(NCI CTCAE v4.0) | Management | Follow up |
| --- | --- | --- |
| Grade 0<br>(i.e. normal / baseline) | Check creatinine at<br>day 7 ± 2 days | If creatinine recheck is abnormal, continue to recheck at<br>least twice a week through the 14-day course of<br>Valacyclovir. |
| Grade 1<br>Creatinine >1 to 1.5 x baseline;<br>>ULN to 1.5 x ULN | Dose level level -1 | If creatinine remains Grade 1 or returns to Grade 0,<br>continue level -1.<br>If creatinine reaches to Grade 2, reduce to level -2.<br>If creatinine reaches Grade 3-4 withhold Valacyclovir. |
| Grade 2<br>Creatinine >1.5 to 3 x baseline;<br>>1.5 to 3 x ULN | Dose level level -2 | If creatinine returns to baseline, continue dose level -1.<br>If creatinine reaches to Grade 2, reduce to level -2.<br>If creatinine reaches Grade 3-4, withhold Valacyclovir. |
| Grade 3<br>Creatinine > 3 x baseline;<br>>3 to 6 x ULN | Withhold<br>Valacyclovir | If creatinine returns to Grade 0, restart at level -1.<br>If creatinine returns to Grade 1, restart at level -2.<br>If creatinine reaches Grade 3 a second time during cycle<br>of Valacyclovir, permanently discontinue unless an<br>associated cause was present (dehydration, concomitant<br>medication) and can be avoided. In such a situation,<br>restart at level -2 and increase to level -1 at discretion of<br>investigator. |
| Grade 4<br>Creatinine >6 x ULN | Withhold<br>Valacyclovir | If creatinine returns to Grade 0, restart at level -1.<br>If creatinine returns to Grade 1, restart at level -2. |

**Table S4.** Patient Demographics and Baseline Characteristics.

| ID | Sex/ Age | KPS | Dual vector dose level | Extent of Surgery #1* | Diagnosis | MGMT promotor methylated | Steroids before surgery #1* | Subsequent surgery: Months from #1 / PD<sup>%tumor if ≤50%</sup> vs PsP<sup>%tumor</sup> |  |  | OS (months) |
| --- | --- | --- | --- | --- | --- | --- | --- | --- | --- | --- | --- |
|  |  |  |  |  |  |  |  | #2 | #3 | #4 |  |
| 1 | F/61-70 | 90 | A | Near GTR | GBM | No | N | 14.5/PD<sup>20%</sup> | 24.0/PD<sup>30%</sup> |  | 27.5 |
| 2 | F/61-70 | 80 | A | GTR | Anaplastic ependymoma | No | Y | 13.0/PD | 24.9/PD | 26.7/PD | 52.7 |
| 3 | F/61-70 | 90 | A | GTR | GBM | Yes | Y | 17.0/PD |  |  | 24.3 |
| 4 | M/71-80 | 90 | B** | Near GTR | GBM | No | Y |  |  |  | ** |
| 5 | M/51-60 | 100 | B | Near GTR | GBM | No | Y | 19.4/PsP<sup>0%</sup> |  |  | 26.1 |
| 6 | F/61-70 | 70 | B | Near GTR | GBM | Yes | N |  |  |  | 5.3 |
| 7 | M/61-70 | 100 | B | Near GTR | GBM | No | Y | 13.9/PsP<sup>2%</sup> |  |  | 21.9 |
| 8 | F/51-60 | 90 | C | Near GTR | GBM | Yes | N | 7.6/PD<sup>45%</sup> | 19.0/PsP<sup>5%</sup> |  | 33.3 |
| 9 | Screen fail (positive pregnancy test) |  |  |  |  |  |  |  |  |  |  |
| 10 | M/61-70 | 70 | C | GTR | GBM | Yes | N |  |  |  | 9.4 |
| 11 | M/61-70 | 90 | C | Near GTR | GBM | No | N |  |  |  | 21.0 |
| 12 | M/41-50 | 100 | D | GTR | Gliosarcoma | No | Y |  |  |  | 11.1 |
| 13 | M/61-70 | 100 | D | GTR | GBM | No | N |  |  |  | 20.5 |
| 14 | M/51-60 | 100 | D | GTR | GBM | No | N | 9.2/PD |  |  | 21.6 |
| 15 | M/71-80 | 100 | E | GTR | GBM | Yes | N |  |  |  | 8.1 |
| 16 | F/61-70 | 90 | E | GTR | GBM | Yes | Y |  |  |  | 57.4+ |
| 17 | Screen fail (frozen pathology: metastatic carcinoma) |  |  |  |  |  |  |  |  |  |  |
| 18 | M/61-70 | 90 | E | GTR | GBM | No | N |  |  |  | 16.6 |
| 19 | M/51-60 | 90 | F | STR | Gliosarcoma | No | Y |  |  |  | 7.1 |
| 20 | F/71-80 | 100 | F | GTR | Gliosarcoma | Yes | Y |  |  |  | 15.7 |
| 21 | F/61-70 | 100 | F | GTR | GBM | No | Y | 16.6/PD<sup>50%</sup> | 28.6/PD<sup>20%</sup> |  | 39.7 |

Abbreviations: KPS, Karnofsky Performance Status; OS, overall survival; GTR, gross total resection; GBM, glioblastoma; PD, progression of disease; PsP, pseudoprogression.
\* Surgery #1 = initial surgery at the time of enrollment.
\*\*Non-evaluable per protocol due to early withdrawal per patient discretion. Dose level subject replaced and subject 4 excluded from survival analysis. See CONSORT flow diagram.

**Table S5.** All Serious Adverse Events.

| Event | Any Grade<br>*no. (%)* | Grade 2-3<br>*no. (%)* | Grade 4-5<br>*no. (%)* |
| --- | --- | --- | --- |
| Gastrointestinal disorders |  |  |  |
| Nausea | 1 (6) | 1 (6) | - |
| General disorders and administration site conditions |  |  |  |
| Pain | 2 (11) | 2 (11) | - |
| Infections and infestations |  |  |  |
| Urinary tract infection | 2 (11) | 2 (11) | - |
| Wound infection | 4 (22) | 4 (22) | - |
| Injury, poisoning and procedural complications |  |  |  |
| Wound dehiscence | 1 (6) | 1 (6) | - |
| Metabolism and nutrition disorders |  |  |  |
| Dehydration | 1 (6) | 1 (6) | - |
| Nervous system disorders |  |  |  |
| Cognitive disturbance | 1 (6) | 1 (6) | - |
| Dysphasia | 2 (11) | 2 (11) | - |
| Edema cerebral | 1 (6) | - | 1 (6) |
| Encephalopathy | 1 (6) | 1 (6)* | - |
| Intracranial hemorrhage | 1 (6) | - | 1 (6) |
| Lethargy | 1 (6) | 1 (6) | - |
| Right side neglect | 1 (6) | 1 (6) | - |
| Seizure | 2 (11) | 2 (11)* | - |
| Syncope | 1 (6) | 1 (6) | - |
| Confusion | 1 (6) | 1 (6) | - |
| Renal and urinary disorders |  |  |  |
| Acute kidney injury | 1 (6) | 1 (6)* | - |
| Respiratory, thoracic and mediastinal disorders |  |  |  |
| Respiratory failure | 1 (6) | - | 1 (6) |
| Vascular disorders |  |  |  |
| Thromboembolic event | 5 (28) | 5 (28) | - |
| Total no. patients with SAE | 10 (56) | 9 (50) | 3 (17) |

\*Possibly attributable to study intervention

**Table S6.** Adverse Events Unrelated to Study Intervention.

| Event | Grade<br>2-3<br>*no. (%)* | Grade<br>4-5<br>*no. (%)* |
| --- | --- | --- |
| Blood and lymphatic system disorders |  |  |
| Anemia | 4 (22) | - |
| Cardiac disorders |  |  |
| Pericardial effusion | 1 (6) | - |
| Ear and labyrinth disorders |  |  |
| Vestibular disorder | 1 (6) | - |
| Gastrointestinal disorders |  |  |
| Abdominal pain | 3 (17) | - |
| Diarrhea | 1 (6) | - |
| Dysphagia | 1 (6) | - |
| Nausea | 3 (17) | - |
| General disorders and administration site conditions |  |  |
| Edema limbs | 2 (11) | - |
| Fatigue | 8 (44) | - |
| Pain | 3 (17) | - |
| Infections and infestations |  |  |
| Upper respiratory infection | 2 (11) | - |
| Urinary tract infection | 3 (17) | - |
| Wound infection | 4 (22) | - |
| Injury, poisoning and procedural complications |  |  |
| Wound dehiscence | 1 (6) | - |
| Investigations |  |  |
| Creatinine increased | 2 (11) | - |
| Neutrophil count decreased | 1 (6) | - |
| Platelet count decreased | 4 (22) | - |
| Metabolism and nutrition disorders |  |  |
| Dehydration | 2 (11) | - |
| Hyperkalemia | 1 (6) | - |
| Hypokalemia | 1 (6) | - |
| Musculoskeletal and connective tissue disorders |  |  |
| Arthralgia | 1 (6) | - |
| Generalized muscle weakness | 2 (11) | - |
| Muscle weakness lower limb | 1 (6) | - |
| Muscle weakness right-sided | 1 (6) | - |
| Pain in extremity | 1 (6) | - |

| Event (cont.) | Grade<br>2-3<br>*no. (%)* | Grade<br>4-5<br>*no. (%)* |
| --- | --- | --- |
| Nervous system disorders |  |  |
| Cognitive disturbance | 1 (6) | - |
| Dysarthria | 2 (11) | - |
| Dysphasia | 3 (17) | - |
| Edema cerebral | - | 3 (17) |
| Facial muscle weakness | 1 (6) | - |
| Headache | 2 (11) | - |
| Intracranial hemorrhage | - | 1 (6) |
| Lethargy | 1 (6) | - |
| Paresthesia | 1 (6) | - |
| Right side neglect | 1 (6) | - |
| Seizure | 2 (11) | - |
| Syncope | 1 (6) | - |
| Visual field deficit | 1 (6) | - |
| Psychiatric disorders |  |  |
| Agitation | 1 (6) | - |
| Confusion | 1 (6) | - |
| Insomnia | 1 (6) | - |
| Respiratory, thoracic and mediastinal disorders |  |  |
| Hypoxia | 1 (6) | - |
| Respiratory failure | - | 1 (6) |
| Skin and subcutaneous disorders |  |  |
| Alopecia | 2 (11) | - |
| Thromboembolic event | 7 (39) | - |

**Table S7.** Antibodies used in Multiplex Immunohistochemistry.

| **Antibody** | **Company** | **Catalog #** | **Clone** | **Dilution** | **Exposure (ms)** |
| --- | --- | --- | --- | --- | --- |
| Anti-Hu CD3e-BX045—Alexa Fluor™ 647 for PhenoCycler | Akoya | 4550119 | AKYP0062 | 1:200 | 250 |
| Anti-Hu CD4-BX003—Alexa Fluor™ 647 for PhenoCycler | Akoya | 4550112 | AKYP0048 | 1:200 | 200 |
| Anti-Hu CD8a-BX026—Atto 550 for PhenoCycler | Akoya | 4250017 | AKYP0028 | 1:200 | 150 |
| Anti-Hu Granzyme B-BX041—Atto 550 for PhenoCycler | Akoya | 4250055 | AKYP0086 | 1:100 | 150 |
| Anti-Hu CD107a-BX006—Alexa Fluor™ 647 for PhenoCycler | Akoya | 4550098 | AKYP0004 | 1:100 | 150 |
| Anti-Hu CD11c-BX024—Alexa Fluor™ 647 for PhenoCycler | Akoya | 4550114 | AKYP0051 | 1:200 | 150 |
| Anti-Hu HLA-DR-BX033—Alexa Fluor™ 647 for PhenoCycler | Akoya | 4550118 | AKYP0063 | 1:200 | 150 |
| Anti-Hu CD45-BX021—Alexa Fluor™ 647 for PhenoCycler | Akoya | 4550121 | AKYP0074 | 1:200 | 150 |
| Anti-Hu SOX2-BX102—Atto 550 for PhenoCycler | Akoya | 4250075 | AKYP0106 | 1:200 | 150 |
| Anti-Hu FOXP3-BX031—Alexa Fluor™ 647 for PhenoCycler | Akoya | 4550071 | AKYP0102 | 1:200 | 250 |
| Anti-Hu IBA1-BX040—Alexa Fluor™ 488 for PhenoCycler | Abcam | Ab221790 | EPR16589 | 1:200 | 150 |

**Supplemental Figure S1.**
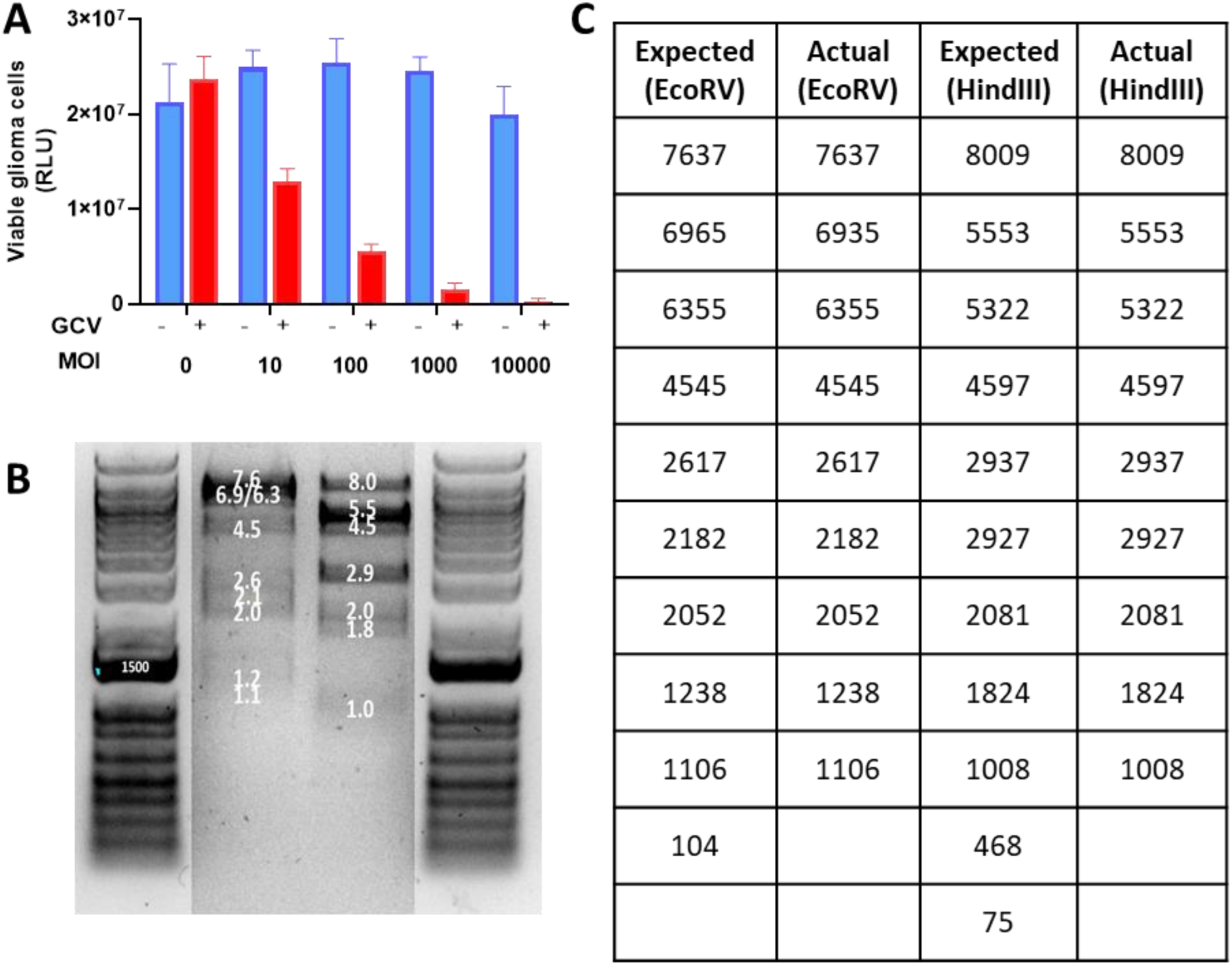
Ad-hCMV-TK expresses bioactive transgene herpes simplex type 1 thymidine kinase (TK). **A)** CNS1 rat glioma cells (2 x 10^4^/well) were infected with Ad-hCMV-TK with 10,000, 1,000, 100, 10, 0 Ad- hCMV-TK/cell. After 24 hours, the cells were incubated for an additional 72 hours in the presence of 1 mg/mL of the prodrug ganciclovir (GCV). Cell viability was determined by CTG assay (Cell Titer-Glo Cell Viability Assays, Promega, Madison, WI). **B)** Restriction Analysis of Ad-hCMV-TK. Viral DNA was isolated from Ad-hCMV-TK, digested with either HindIII or EcoRV, and run on a 0.8% agarose gel. Image of gel electrophoresis displays a gel after 90 min of electrophoresis. **C)** Table comparing the predicted and actual DNA size following restriction analysis. All sizes are described in base pairs (bp).

**Supplemental Figure S2.**
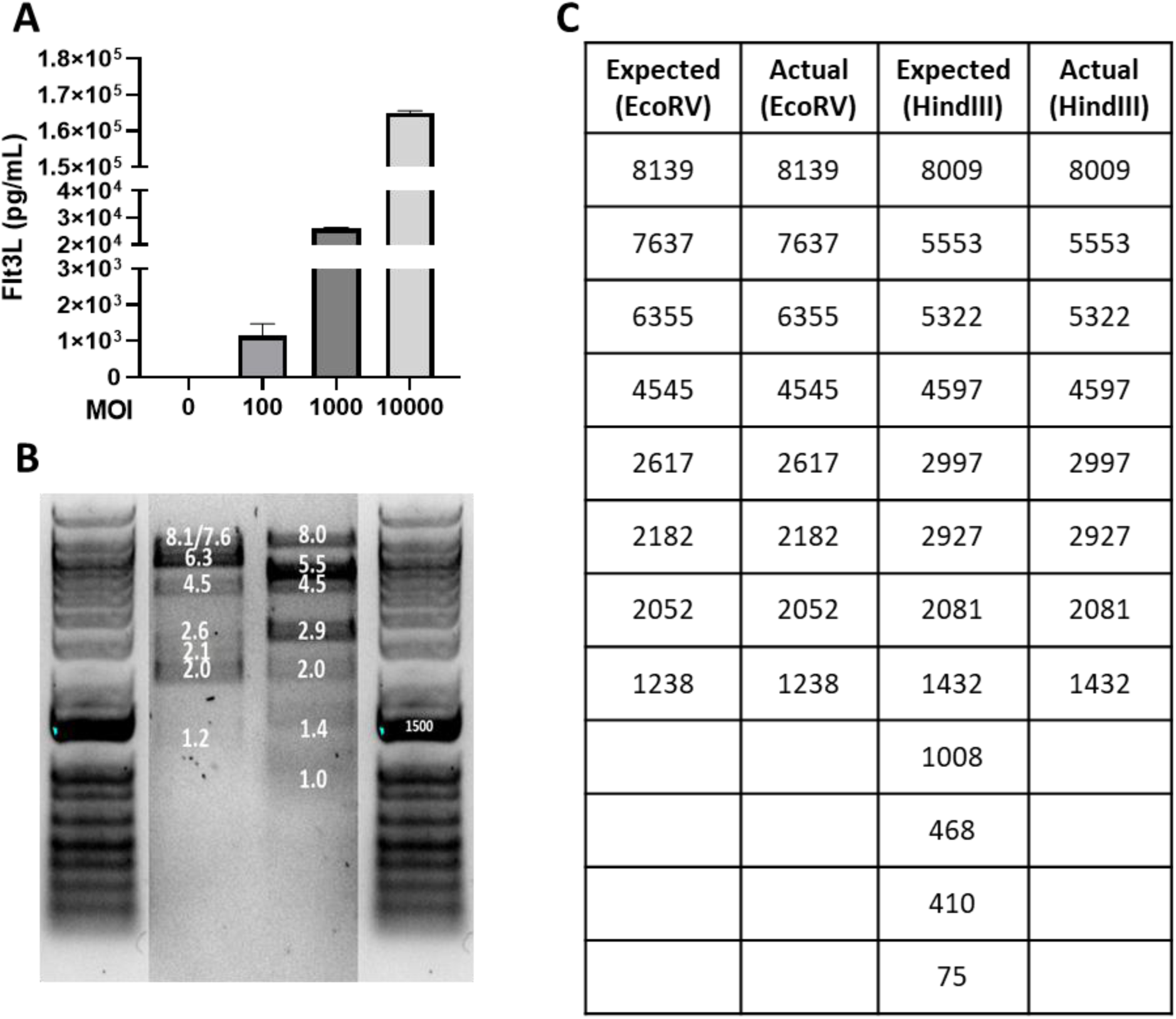
Ad-hCMV-Flt3L expresses its transgene human soluble FMS-like tyrosine kinase 3 ligand. **A)** CNS1 rat glioma cells (2 x 10^4^well) were infected with Ad-hCMV-Fl3tL with 10,000, 1,000, 100, 10, 0 Ad- hCMV-Fl3tL /cell. After 72 hours, the levels of hFlt3L were measured in the cell supernatants using Flt3L specific ELISA. **B)** Restriction Analysis of Ad-hCMV-TK. Viral DNA was isolated from Ad-hCMV-Flt3L, digested with either HindIII or EcoRV, and run on a 0.8% agarose gel. Image of gel electrophoresis displays a gel after 90 min of electrophoresis. **C)** Table comparing the predicted and actual DNA size following restriction analysis. All sizes are described in base pairs (bp).

**Supplemental Figure S3.**
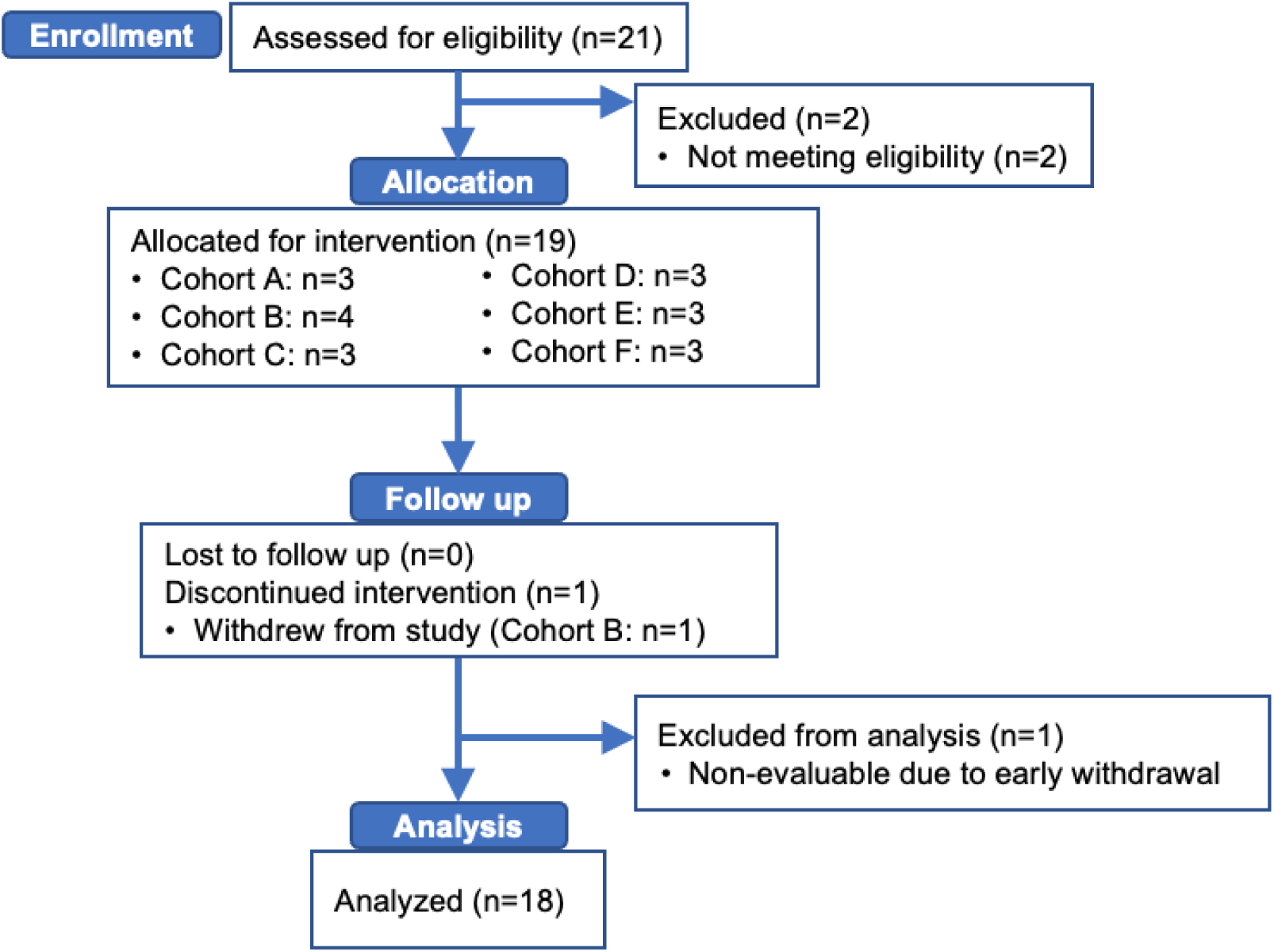
The consort flow diagram. Flow diagram of the enrollment, intervention allocation, follow-up, and data analysis of the dose-finding trial of combined cytotoxic and immune-stimulatory and adenovirally mediated strategy using Flt3L and HSV1-TK.

**Supplemental Figure S4.**
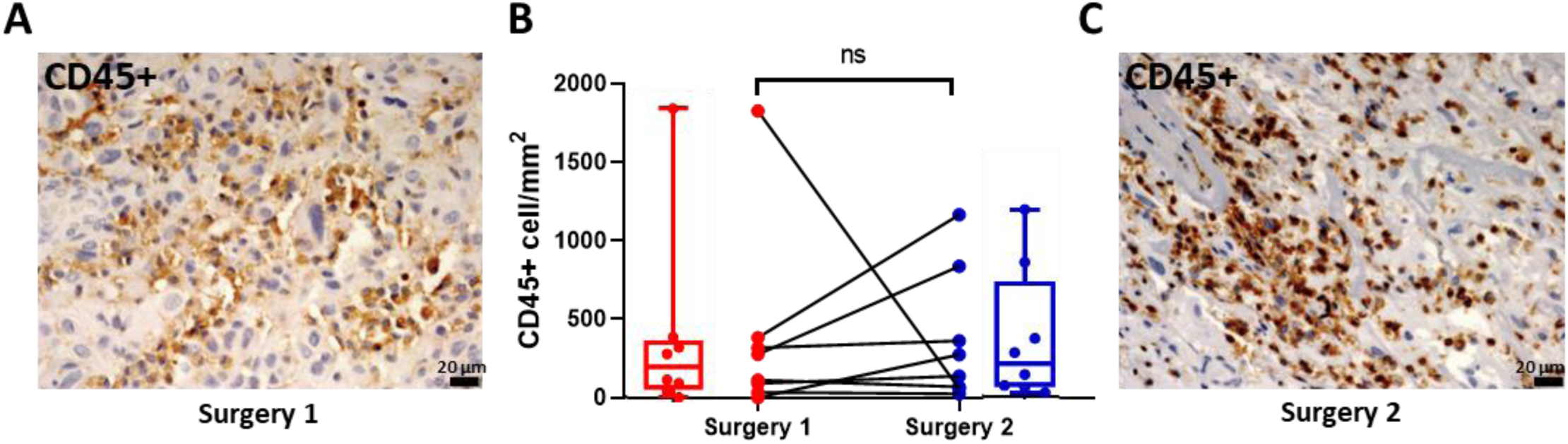
Immunohistochemical analysis of CD45+ cells in human glioblastoma tissue at primary resection (surgery 1), pre vector TK-Flt3L treatment, and at secondary surgery due to tumor recurrence. **A and C)** Representative images of CD45 expression of patient ID #1. Scale bar: 20μm. **B)** Box plots and lines graphs represent the quantification of CD45+ cells numbers (cells/mm2) using QuPath positive cell detection on glioblastomas samples obtained pre-TK-Flt3L treatment (Surgery 1) and post-TK-Flt3L treatment (Surgery 2). Error bars represent ±SEM. n=8 patients, ns = non-significant. For each marker, a linear mixed effects (LME) model was used to compare NumPos (log-transformed) between two surgery procedures. Random effect was included in the LME model to consider correlations between data measured on the same subject. Significance is determined if p<0.05. All analyses were conducted using SAS (version 9.4, SAS Institute, Cary, NC).

**Supplemental Figure S5.**
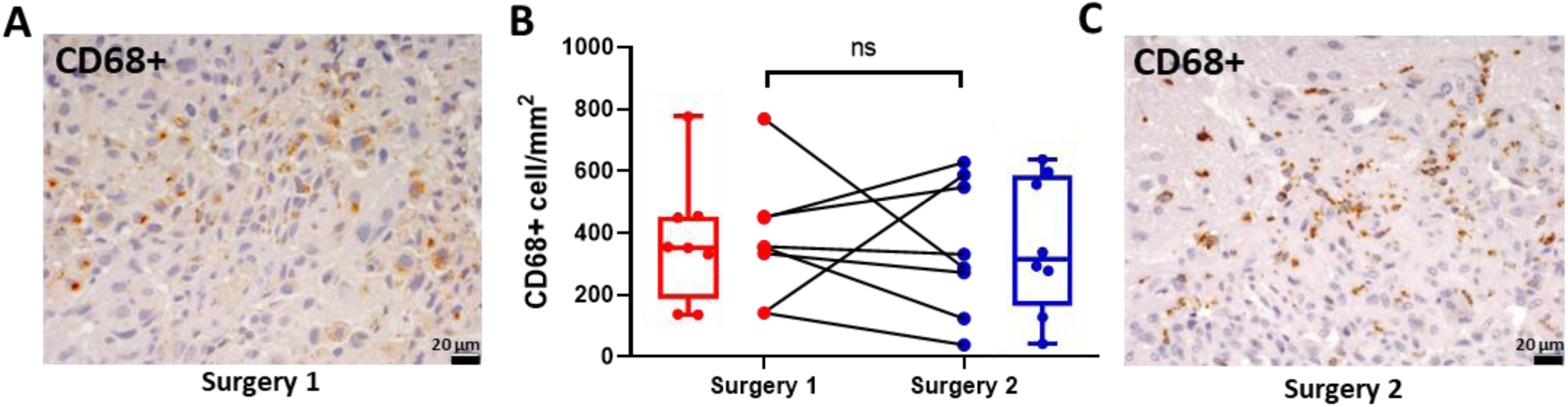
Immunohistochemical analysis of CD68+ cells in human glioblastoma tissue at primary resection (surgery 1), pre vector TK-Flt3L treatment, and at secondary surgery due to tumor recurrence. **A and C)** Representative images of CD68 expression of patient ID #1. Scale bar: 20μm. **B)** Box plots and lines graphs represent the quantification of CD68+ cells numbers (cells/mm2) using QuPath positive cell detection on glioblastomas samples obtained pre-TK-Flt3L treatment (Surgery 1) and post-TK-Flt3L treatment (Surgery 2). Error bars represent ±SEM. n=8 patients, ns = non-significant. For each marker, a linear mixed effects (LME) model was used to compare NumPos (log-transformed) between two surgery procedures. Random effect was included in the LME model to consider correlations between data measured on the same subject. Significance is determined if p<0.05. All analyses were conducted using SAS (version 9.4, SAS Institute, Cary, NC).

**Supplemental Figure S6.**
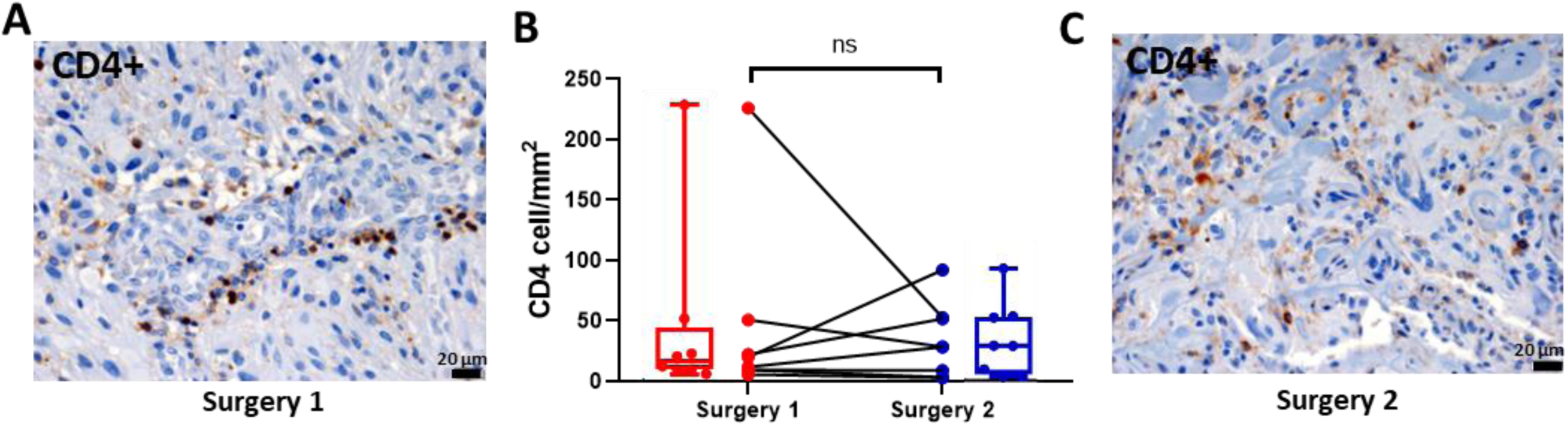
Immunohistochemical analysis of CD4+ cells in human glioblastoma tissue at primary resection (surgery 1), pre vector TK-Flt3L treatment, and at secondary surgery due to tumor recurrence. **A and C)** Representative images of CD4 expression of patient ID #1. Scale bar: 20μm. **B)** Box plots and lines graphs represent the quantification of CD4+ cells numbers (cells/mm2) using QuPath positive cell detection on glioblastomas samples obtained pre-TK-Flt3L treatment (Surgery 1) and post-TK-Flt3L treatment (Surgery 2). Error bars represent ±SEM. n=8 patients, ns = non-significant. For each marker, a linear mixed effects (LME) model was used to compare NumPos (log-transformed) between two surgery procedures. Random effect was included in the LME model to consider correlations between data measured on the same subject. Significance is determined if p<0.05. All analyses were conducted using SAS (version 9.4, SAS Institute, Cary, NC).

**Figure S7:**
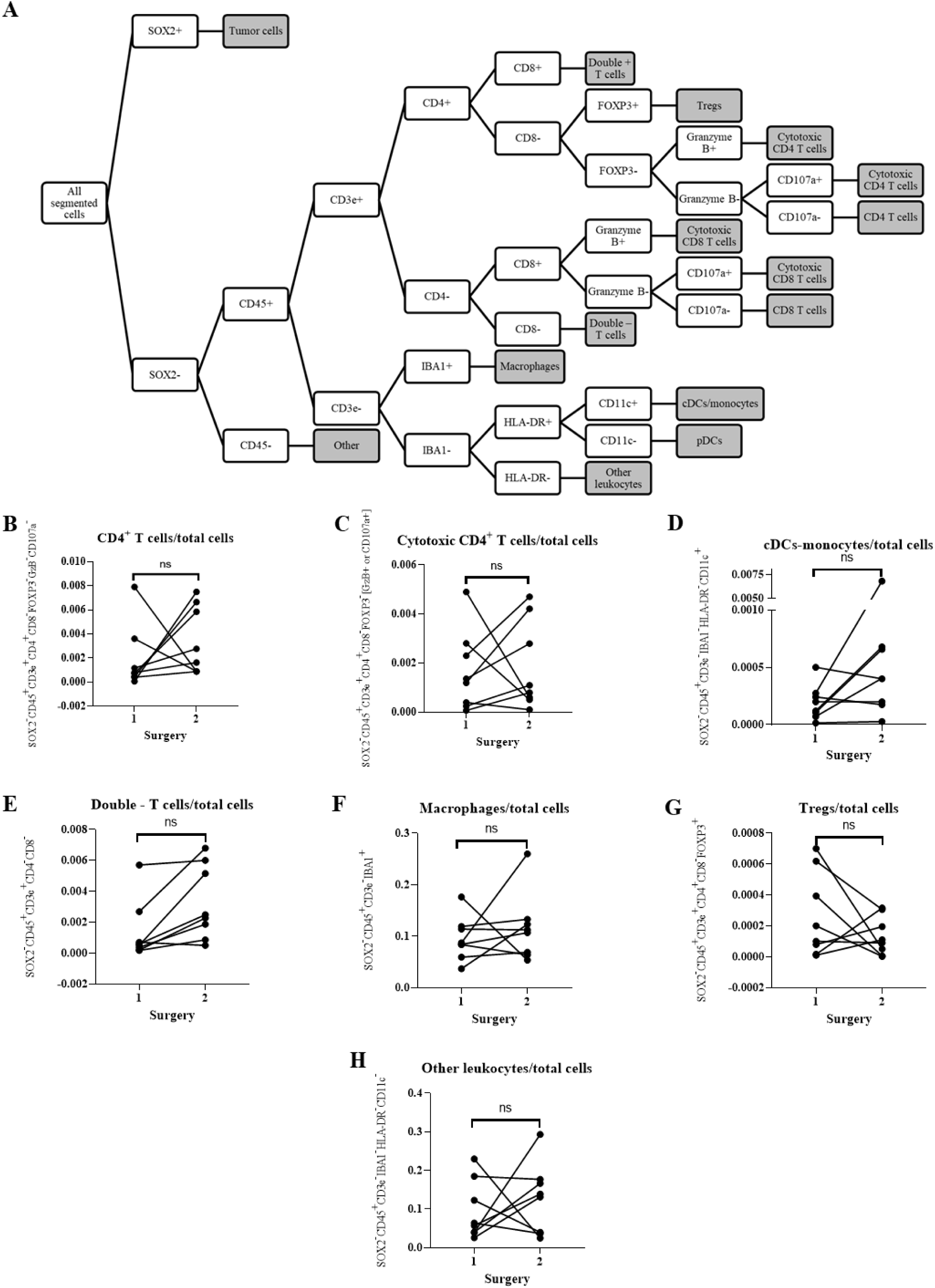
Phenotyping and quantification of immune cell types in the biopsies obtained at primary tumor resection and post dual-vector TK-Flt3L treatment at tumor recurrence. Cell classification was performed using a decision tree shown with single intensity thresholds for each marker, coded into a script used in QuPath **(A)**. Spaghetti plots were generated to represent the percentage of CD4+ T cells out of total cells **(B)**, cytotoxic CD4+ T cells out of total cells **(C)**, cDCs/monocytes out of total cells **(D)**, double negative T cells out of total cells **(E)**, macrophages out of total cells **(F)**, Tregs out of total cells **(G)**, and other leukocytes out of total cells **(H)** in surgery 1 and surgery 2 samples. A generalized linear mixed effects regression model was employed to compare the rates of cell type counts between two different time points. The model assumed a negative binomial distribution, with a log link function and log (total cell) included as the offset in the regression. The results reported included the rate ratio (RR) and p-value. To control the family-wise type I error rate, the p-values were adjusted using the Sidak correction method.

**Figure S8:**
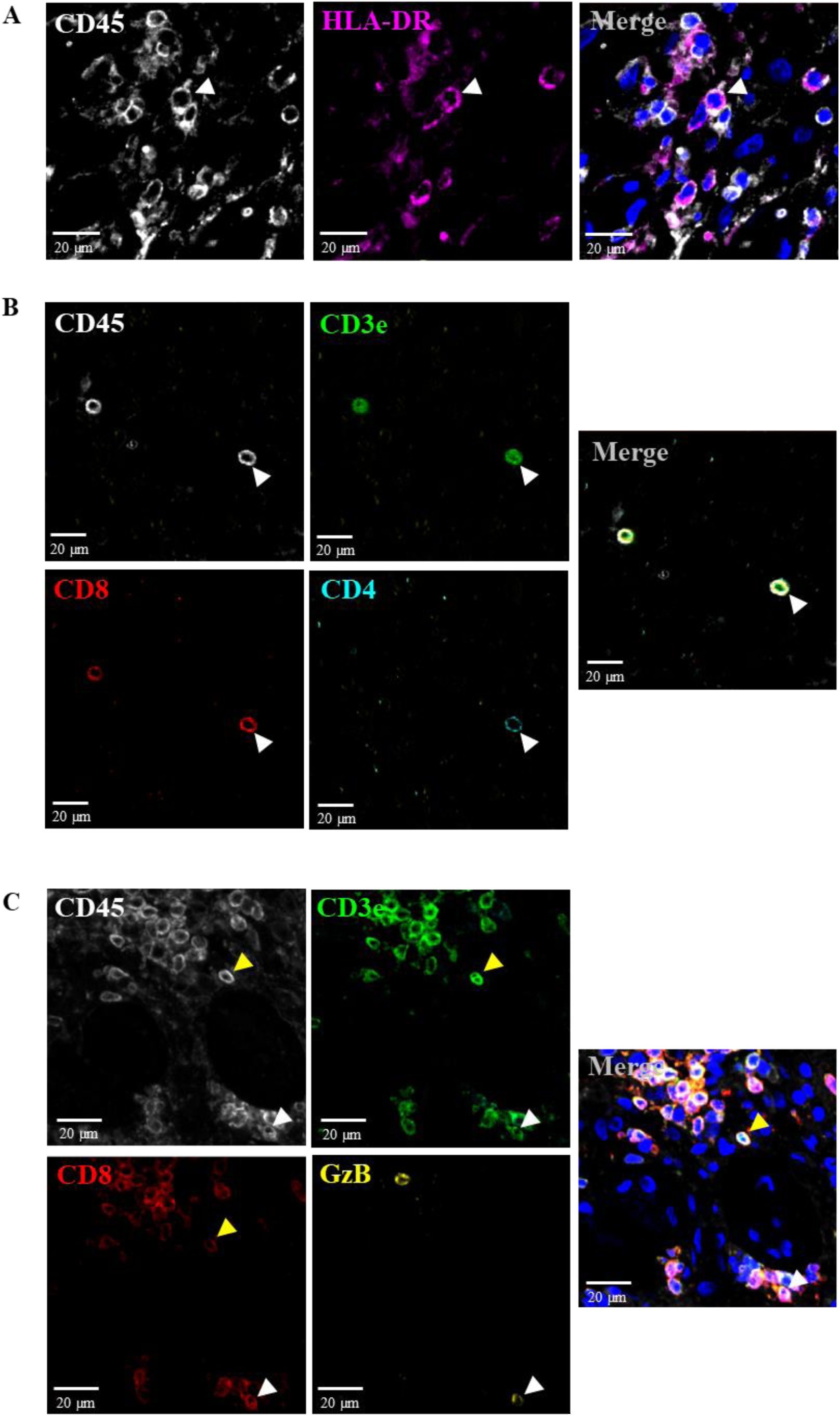
Multiplex immunocytochemical analysis of immune cells in the biopsies obtained post-dual vector TK- Flt3L treatment at tumor recurrence. Panel A) shows representative images of colocalization of CD45 and HLA-DR expression, which were used to identify pDCs (Patient ID #1; surgery 2). In panel B), representative images of colocalizing CD45, CD3e, CD8, and CD4 expression were presented, which were used to identify double positive T cells (Patient ID #7; surgery 2). Panel C) displays representative images of colocalizing CD45, CD3e, CD8, and granzyme B expression, which were used to identify both cytotoxic CD8+ T cells and CD8+ T cells (Patient ID #1; surgery 2).

**Figure S9:**
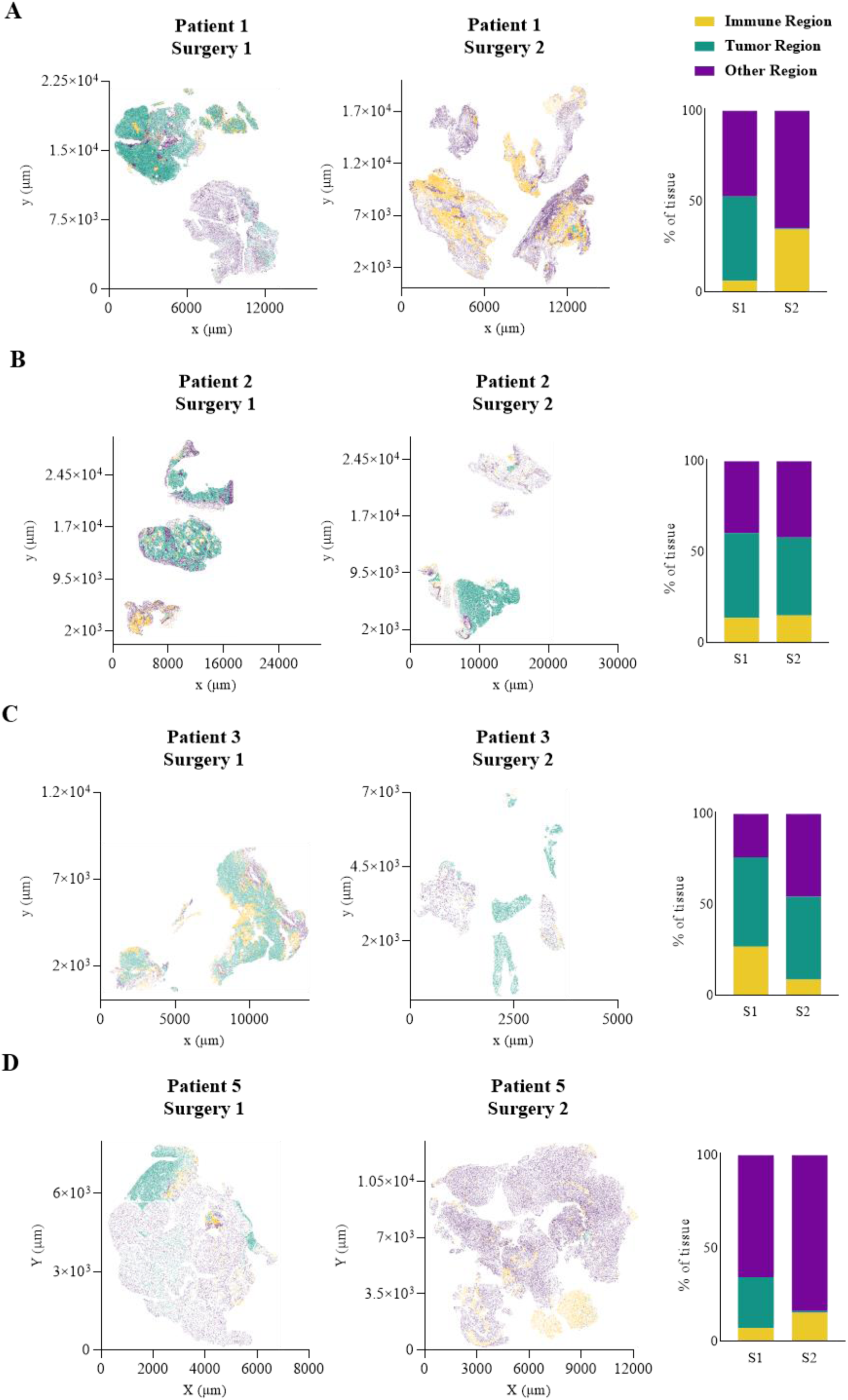
Neighborhood analysis of the tumor microenvironment in the biopsies obtained at primary tumor resection and at tumor recurrence. The plots illustrate the cells present in the resected tissue of both primary (on the left) and recurrent (in the middle) tumors, with each cell colored according to its corresponding region type for the specified patient. The accompanying bar graph on the right displays the prevalence of each region type in the tissue samples. The four different patients are labelled as follows: **A)** patient 1, **B)** patient 2, **C)** patient 3, and **D)** patient 5.

**Figure S10:**
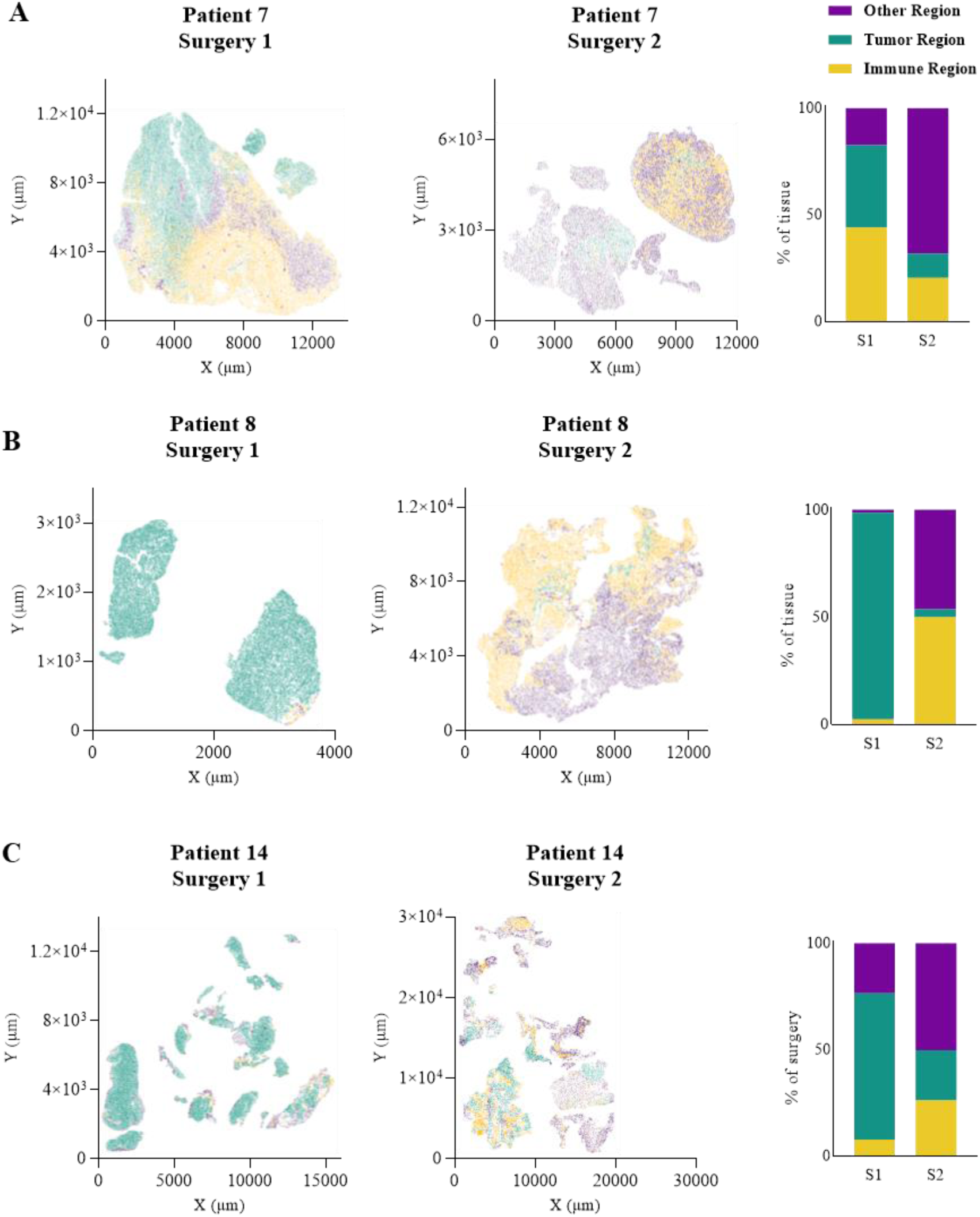
Neighborhood analysis of the tumor microenvironment in glioblastoma tissues at primary tumor resection and at tumor recurrence. The figures depict the cellular composition of resected tissue from primary (on the left) and recurrent (in the middle) tumors, with each cell colored based on its corresponding region type for the indicated patient. The corresponding bar graph on the right illustrates the relative prevalence of each region type detected in the tissue specimens. The three patients are labelled as follows: **A)** patient 7, **B)** patient 8, and **C)** patient 14.

**Figure S11.**
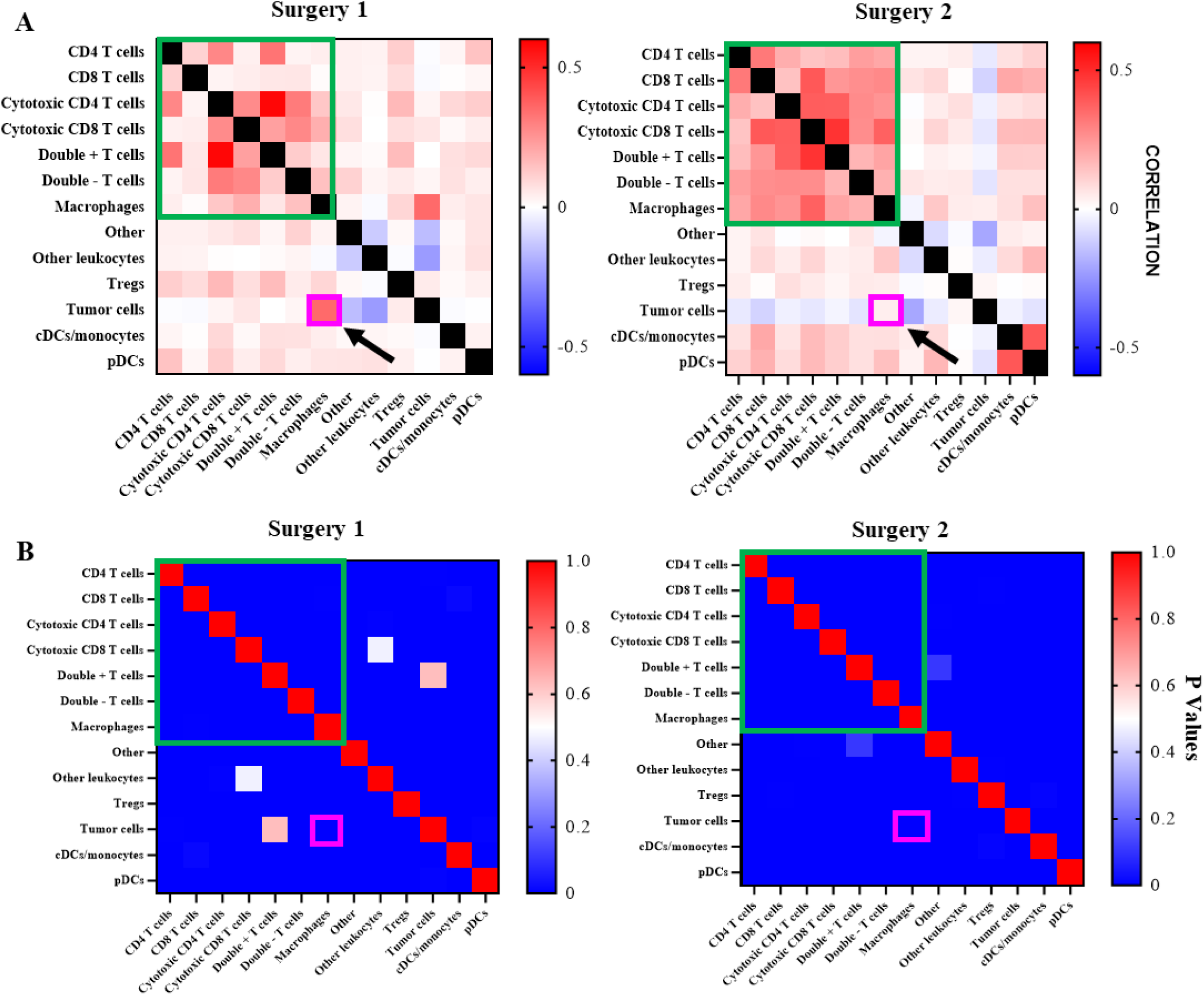
Comparison of average cell-cell correlations across six patients exhibiting immune infiltration at tumor recurrence. **A)** Average cell-cell correlations were measured using Pearson’s correlation coefficient, for patients 1, 5, 7, 8, 14 & 21 at primary tumor resection (Surgery 1) and up on tumor recurrence (Surgery 2). Interactions between T cells and macrophages denoted with green box, while cell-cell association between macrophages and tumor cells denoted with pink box (indicated by black arrows). The green box denotes interactions between T cells and macrophages, and the higher cell-cell associations between many T cells and macrophages at surgery 2 indicate increased interactions between these immune cell types. Decreased cell-cell correlation between macrophages and tumor cells compared to surgery 1 is denoted with a pink box. **B)** Heat maps of p-values corresponding to correlations in panel A.

**Figure S12:**
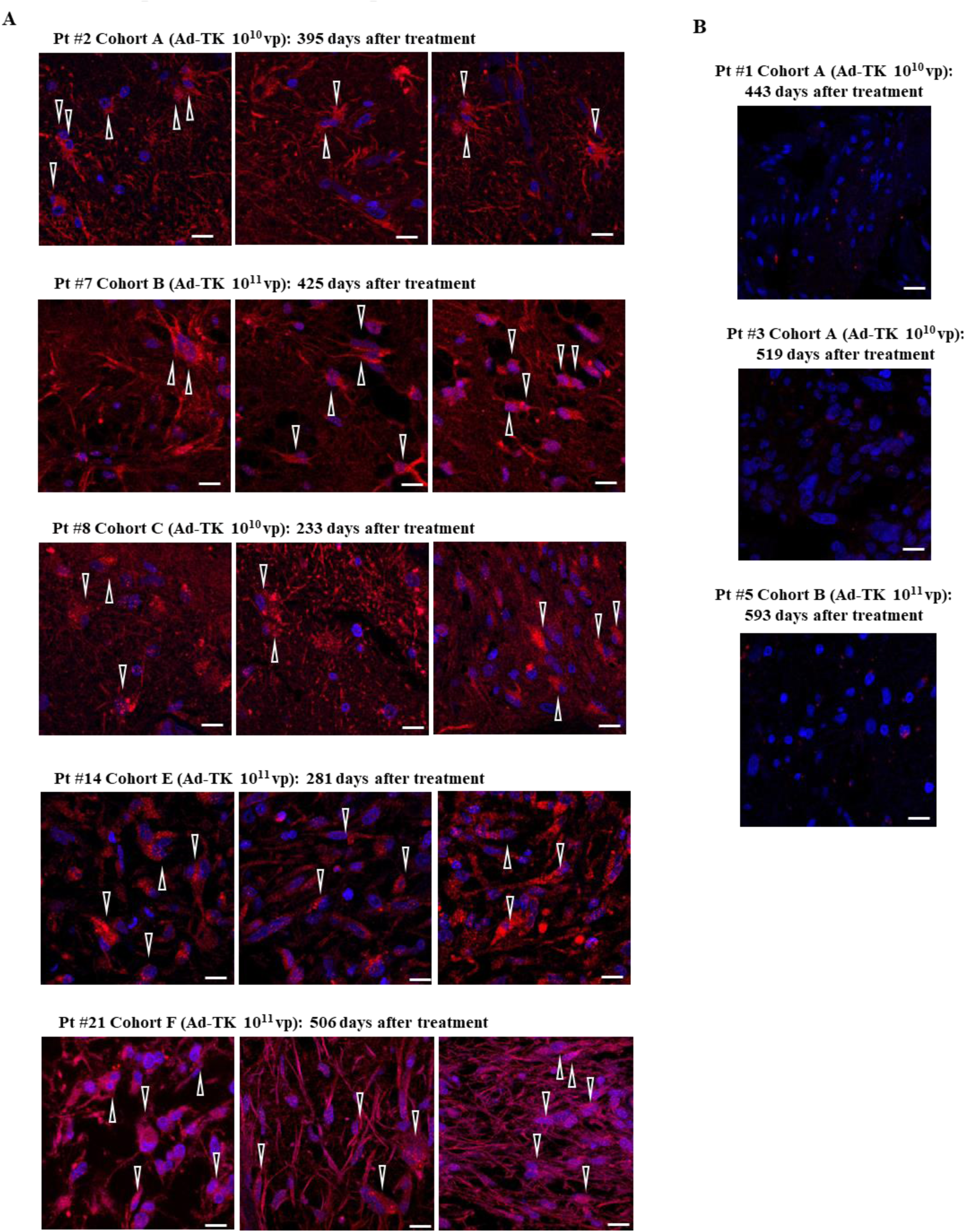
Persistent expression of HSV1-TK post dual-vector TK-Flt3L treatment at tumor recurrence. To assess the long-term expression of HSV1-TK (red), we utilized sections of recurrent tumor biopsies obtained at second surgery. **A)** The representative sections of patients who tested positive for HSV1-TK are displayed, along with information about the viral dosage received and the number of days that elapsed after treatment administration. The nuclei were counterstained with DAPI (blue). We could detect HSV1-TK in 5 out of 8 samples from surgery 2, up to 17 months post-viral administration. B). Representative sections of patients who tested negative for HSV1-TK are displayed. All the scale bars are of 10 µm. Arrows point to highly reactive cells.

**Figure S13:**
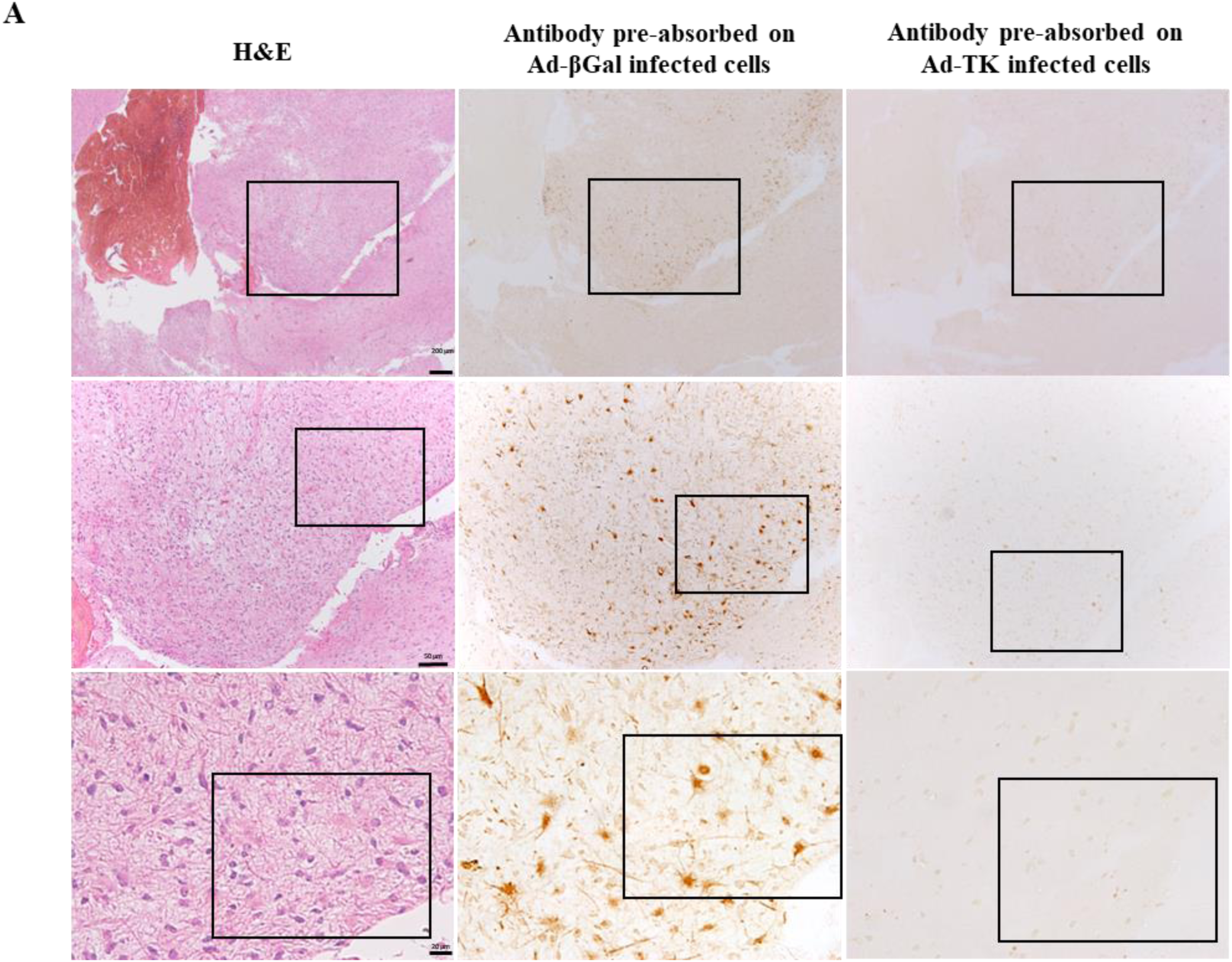
Neutralization experiments to validate the specificity of HSV1-TK detection. **A)** The consecutive sections of surgery 2 (recurrent tumor) from patient #21 were used in the same location to demonstrate the specificity of the HSV1-TK antibody. The antibody was pre-incubated with cells infected with either Ad-βGal or Ad-TK. Pre-absorption with an unspecific protein (βGal) did not interfere with the positive cells, whereas pre-absorption with HSV1-TK neutralized the antibody, thereby demonstrating the specificity of this detection. Hematoxylin and eosin staining of the same area was imaged to provide a general overview of the tissue at that location. Each square denotes the inset displayed in the image below, with the highest magnification displayed in the main manuscript Figure 5B.

**Figure S14:**
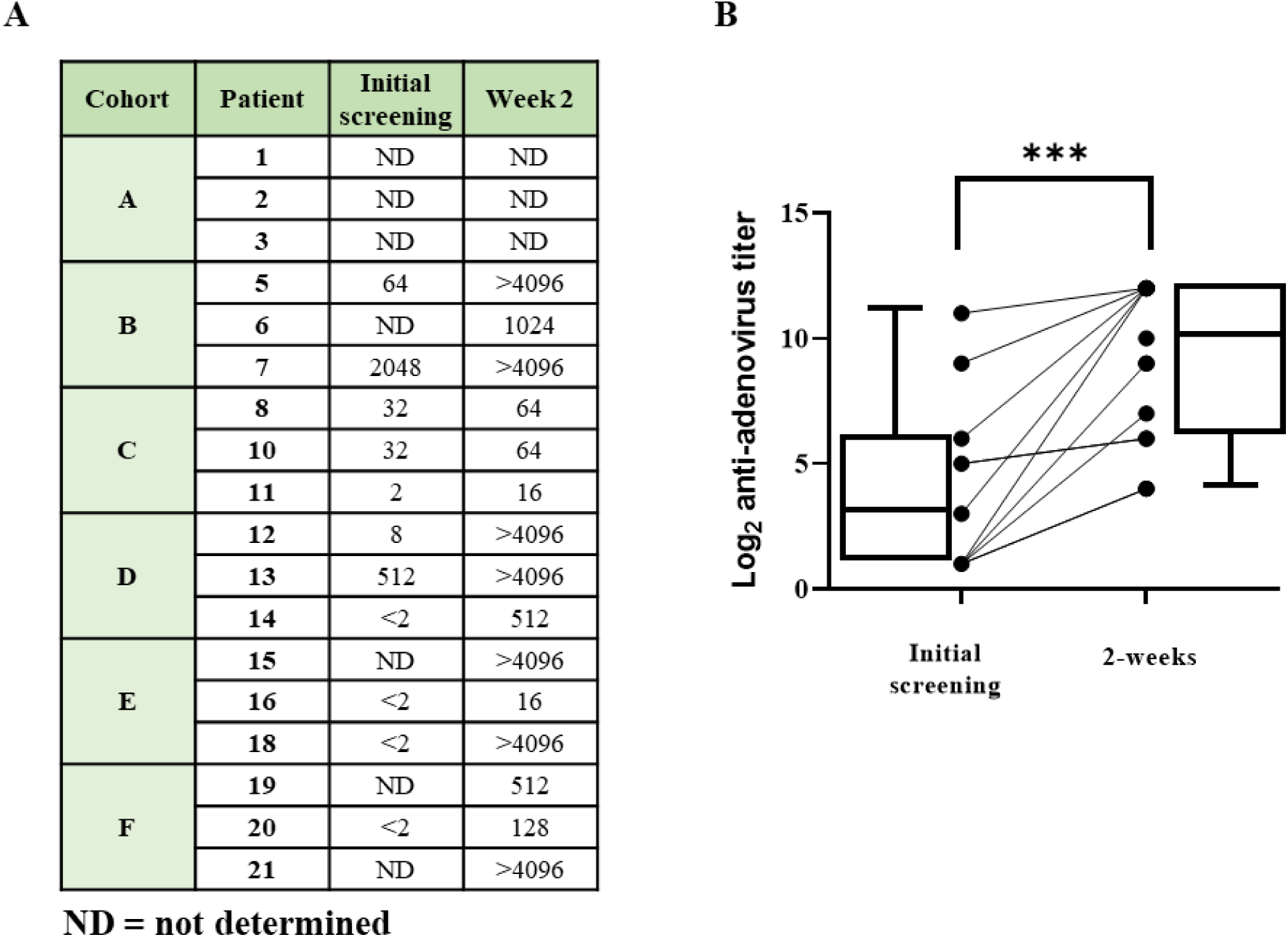
Detection of anti-adenovirus antibodies in plasma following dual vector treatment. **A)** Cohort B-F patients’ plasma samples were collected during initial screening (before surgery) and again 2 weeks after surgery/treatment. These samples were then tested for anti-adenovirus antibodies using a neutralization assay, with the titer indicating the level of antibodies present - the higher the number, the stronger the anti-adenoviral response. **B)** The results were plotted to show the anti-adenovirus antibody titer (log2) at the initial screening and 2 weeks after treatment administration. The statistical analysis revealed a significant difference, with a p-value of 0.0005.

**Figure S15:**
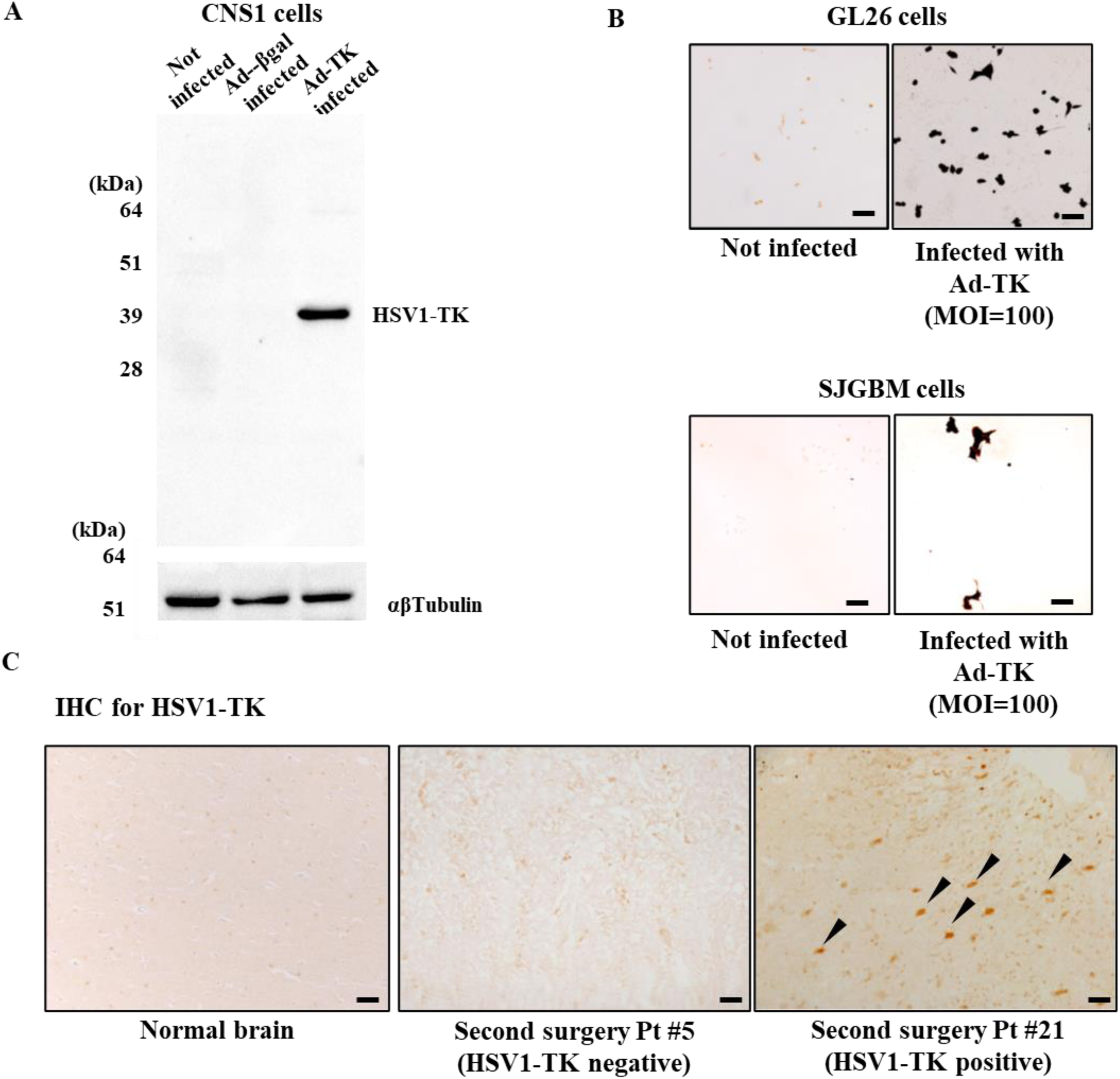
HSV1-TK antibody validation. **A)** Rat glioma cells CNS1 were: (1) not infected (control), (2) infected Ad-βGal (MOI=50), or (3) infected with Ad-TK (MOI=50). After treatment cells were lysed, and 20µg of protein per sample was subjected to SDS-PAGE followed by Western Blot. Membranes were incubated with polyclonal anti-HSV1- TK (1:1000). Immunoreactive band size is 41k Da, this being the expected size of HSV1-TK. Notice no non-specific bands were detected in the control or in the Ad-βGal infected samples. Anti-αβTubulin (1:3000) was used as a loading control. **B)** Mouse glioma cells GL26 and human glioma cells SJGBM were infected with Ad-TK (MOI=100). Forty-eight hours after infection cells were fixed and subjected to immunocytochemistry, employing polyclonal anti-HSV1- TK (1:1000). Both cell lines displayed a strong immunoreactivity post infection, while having none in the control condition. **C)** Normal brain tissue and samples from surgeries 2 were subjected to immunohistochemistry with polyclonal anti-HSV1-TK (1:1000). Normal brain had no immunostaining (left). Five out of 8 patients had HSV1-TK positive immunoreactivity. As examples, patient #5 was negative (center) and patient #21 was positive (right). Scale bar is 50µm.

